# Smartphone-based digital biomarkers for Parkinson’s disease in a remotely-administered setting

**DOI:** 10.1101/2021.01.13.21249660

**Authors:** María Goñi, Simon B. Eickhoff, Mehran Sahandi Far, Kaustubh R. Patil, Juergen Dukart

## Abstract

Smartphone-based digital biomarker (DB) assessments provide objective measures of daily-life tasks and thus hold the promise to improve diagnosis and monitoring of Parkinson’s disease (PD). To date, little is known about which tasks perform best for these purposes and how different confounds including comorbidities, age and sex affect their accuracy. Here we systematically assess the ability of common self-administered smartphone-based tasks to differentiate PD patients and healthy controls (HC) with and without accounting for the above confounds. Using a large cohort of PD patients and healthy volunteers acquired in the mPower study, we extracted about 700 features commonly reported in previous PD studies for gait, balance, voice and tapping tasks. We perform a series of experiments systematically assessing the effects of age, sex and comorbidities on the accuracy of the above tasks for differentiation of PD patients and HC using several machine learning algorithms. When accounting for age, sex and comorbidities, the highest balanced accuracy on hold-out data (73%) was achieved using random forest when combining all tasks followed by tapping using relevance vector machine (67%). Only moderate accuracies were achieved for other tasks (60% for balance, 56% for gait and 53% for voice data). Not accounting for the confounders consistently yielded higher accuracies of up to 77% when combining all tasks. Our results demonstrate the importance of controlling DB data for age and comorbidities.

## I. INTRODUCTION

Diagnosis of Parkinson’s disease (PD) still often relies on in-clinic visits and evaluation based on clinical judgement as well as patient and caregiver reported information. This lack of objective measures and the need for in-clinic visits result in the often late and initially inaccurate diagnosis [1]. Recent studies have identified digital assessments as such promising objective biomarkers for PD symptoms including bradykinesia [2], [3], freezing of gait [4], [5], impaired dexterity [6], balance and speech difficulties [7]–[9]. Most of these results were obtained with a moderate number of participants and in a standardized and controlled clinical setting, reducing generalizability and limiting an interpretation with respect to applicability of these measures to an at-home self-administered setting [10]–[12].

As most relevant sensors deployed in these in-clinic studies are also embedded in modern smartphones, this opens the possibility to collect such objective, reliable and quantitative information as digital biomarkers (DB) in an at-home setting and therewith to facilitate diagnosis, health monitoring or treatment management using low-cost, simple and portable technology [13]. Indeed, recent studies applying machine learning algorithms to these high-dimensional data suggested a good diagnostic sensitivity of the respective digital assessments for detection of Parkinson’s disease [14]–[17]. However, such at-home assessments create a range of new challenges including selection bias, confounding and sources of noise that need to be understood and dealt with to ensure good reliability of respective outcomes to a level that is sufficient for at home data collection [18]. For example, age, sex and comorbidities are known confounding factors that impact many measures of disease symptoms across neurodegenerative diseases including PD [19]–[23]. Yet, several studies eluded the importance of matching and controlling for these variables [24]–[26], including age, sex [27], [28] or comorbidities which might induce motor (i.e. bradykinesia, tremor or rigidity) and non-motor (i.e. fatigue, restless legs or sleep) symptoms [25]. Other potential data collection biases include small sample sizes [14], [29], inclusion of several recordings per subject [15], [27] or signals of different time lengths [28], which may potentially lead the classifier to detect the idiosyncrasies of each subject rather than specific PD related symptoms, as demonstrated by Neto et al. [30]–[32]. In addition, replicability of results is rarely performed in current studies, which may lead to lack of generalizability. Despite the considerable promise for DB in healthcare, these issues limit comparability across studies, hindering interpretation and obstructing translation to the clinic.

Recently, a large dataset of at-home smartphone-based assessments of commonly applied PD tasks including gait, balance, finger tapping and voice evaluations was collected in the mPower study providing a unique resource to examine DB in the study of PD [33], [34]. Indeed, several studies applying machine learning (ML) algorithms have employed this dataset in the study of PD diagnosis, achieving quite different results across studies. Whilst plausible, the impact of the aforementioned confounds on ML-based detection of PD using different at-home digital assessments has not been yet systematically established and has indeed been ignored in many previous studies [15], [24], [28], [35], [36].

Here we systematically explore the influence of accounting for age, sex and comorbidities in the detection of PD in a large at-home dataset. Concretely, we use the mPower dataset to evaluate the ability of common DB task (gait, balance, voice, tapping) for differentiation between PD and HC. In addition, we identify potential DB of Parkinson’s disease. With this work, we aim to outline practical suggestions to guide future studies practices and improve comparability across studies.

## II. METHODS

### A. DATA

Data used in this work were derived from the mPower study [33]. MPower is a mobile application-based study to monitor indicators of PD progression and diagnosis by the collection of data in subjects with and without PD. Using this app, subjects were presented with a one-time demographic survey about general demographic topics and health history. Completion of the Movement Disorder Society’s Unified Parkinson’s Disease Rating Scale (MDS-UPDRS) and the Parkinson’s Disease Questionnaire short form (PDQ-8) surveys used for PD assessment was requested at baseline as well as monthly throughout the course of the study. Due to the length of the MDS-UPDRS instrument, subjects were presented only a subset of questions focusing largely on the monitor symptoms of PD [33]. Participants had to select “true” or “false” to the following question “Have you been diagnosed by a medical professional with Parkinson Diseaseã”. According to this answer, they were classified as Parkinson’s Disease (PD) or Healthy Control (HC). Subjects who did not answer this question were discarded from further analysis. All subjects were presented with different tasks including gait, balance, voice and tapping, which they could complete up to 3 times per day. Subjects who self-identified as having a professional diagnosis of PD were asked to perform these tasks (1) immediately before taking their medication, (2) after taking their medication and (3) at some other time (Table 8). Subjects who self-identified as not having a diagnosis of PD could complete these tasks at any time during the day. In the gait task, subjects were asked to walk 20 steps in a straight line. In the balance task they were required to stand still for 30 seconds. During the voice activity task, subjects were requested to say ‘Aaah’ into the microphone for 10 seconds. Finally, during the tapping task participants were instructed to alternatively tap two points on the screen within a 20 seconds interval. We additionally excluded those subjects who gave no information about their age, sex or had inconsistencies in their clinical data (e.g. self-reported healthy controls who answered questions about PD diagnosis or PD medication). Since the mPower dataset is strongly slanted toward young HC (Table 15), we restricted our analysis to those subjects within the age range of 35 to 75 years old. This cleaning step resulted in the exclusion of 40-50% of the data depending on the task. To avoid “learning effects” and biases due to several recordings, we only considered the first recording of each subject in the analyses. Further details about data cleaning can be found in Appendix I. Demographic details are shown in Table 1.

**Table 1.** Demographics for PD and HC subjects for each experiment. Those cases where age or sex are significantly different between PD and HC are indicated with an asterisk (2 sample t-test for age and Chi-square for sex with 95% confidence)

|  | **Gait** |  | **Balance** |  | **Voice** |  | **Tapping** |  | **Multimodal** |  |
| --- | --- | --- | --- | --- | --- | --- | --- | --- | --- | --- |
|  | PD | HC | PD | HC | PD | HC | PD | HC | PD | HC |
| **Experiment 1 (all)** |  |  |  |  |  |  |  |  |  |  |
| N | 610 | 787 | 612 | 803 | 893 | 1257 | 970 | 1630 | 597 | 742 |
| Male/ | 399/ | 640/ | 401/ | 653/ | 571/ | 1018/ | 630/ | 1336/ | 390/ | 607/ |
| female | 211* | 147* | 211* | 150* | 322* | 239* | 340* | 294* | 207* | 135* |
| Age (mean±sd) | 60.3 ± 8.94* | 49.04 ± 10.71* | 60.29 ± 8.94* | 48.9 ± 10.72* | 60.13 ± 8.97* | 47.65 ± 10.41* | 59.85 ± 9.05* | 46.84 ± 10.05* | 60.36 ± 8.86* | 49.22 ± 10.78* |
| UPDRS | 13.17 ± |  | 13.14 ± |  | 13.48 ± |  | 13.44 ± |  | 13.15 ± 7.8 |  |
| mean±sd (n) | 7.78 (350) | - | 7.78 (351) | - | 7.93 (566) | - | 7.89 (588) | - | (344) | - |
| UPDRS I | 5.66 ± 3.64 | - | 5.64 ± 3.64 | - | 5.64 ± 3.63 | - | 5.63 ± 3.63 | - | 5.64 ± 3.64 | - |
| mean±sd (n) | (361) | - | (362) | - | (586) | - | (608) | - | (355) | - |
| UPDRS II | 7.59 ± 5.18 | - | 7.58 ± 5.18 | - | 7.9 ± 5.53 | - | 7.86 ± 5.49 | - | 7.59 ± 5.21 | - |
| mean±sd (n) | (350) | - | (351) | - | (572) | - | (594) | - | (344) | - |
| **Experiment 2 (matched)** |  |  |  |  |  |  |  |  |  |  |
| N | 373 | 373 | 376 | 376 | 534 | 534 | 608 | 608 | 361 | 361 |
| Male/ | 278/ | 286/ | 280/ | 288/ | 379/ | 394/ | 435/ | 450/ | 270/ | 278/ |
| female | 95 | 87 | 96 | 88 | 155 | 140 | 173 | 158 | 91 | 83 |
| Age (mean±sd) | 57.09 ± 9.4 | 57.09 ± 9.4 | 57.1 ± 9.4 | 57.1 ± 9.4 | 56.54 ± 9.3 | 56.54 ± 9.3 | 56.38 ± 9.23 | 56.38 ± 9.23 | 57.18 ± 9.36 | 57.18 ± 9.36 |
| UPDRS | 14.38 ± |  | 13.61 ± |  | 13.76 ± |  | 13.7 ± 8.08 |  | 14.37 ± |  |
| mean±sd (n) | 8.48 (206) | - | 8.34 (202) | - | 8.21 (324) | - | (349) | - | 8.51 (190) | - |
| UPDRS I | 6.16 ± 3.91 | - | 5.73 ± 3.85 | - | 5.84 ± 3.78 | - | 7.8 ± 3.71 | - | 5.99 ± 3.82 | - |
| mean±sd (n) | (213) | - | (207) | - | (333) | - | (361) | - | (198) | - |
| UPDRS II | 8.27 ± 5.66 | - | 7.95 ± 5.49 | - | 7.98 ± 5.6 | - | 7.97 ± 5.53 | - | 8.45 ± 5.69 | - |
| mean±sd (n) | (206) | - | (202) | - | (328) | - | (352) | - | (190) | - |
| **Experiment 3 (no comorbidities, matched), experiment 4 (no comorbidities, matched, age controlled), experiment 5 (no comorbidities, matched, sex controlled) and experiment 6 (no comorbidities, matched, controlled)** |  |  |  |  |  |  |  |  |  |  |
| N | 317 | 317 | 320 | 320 | 446 | 446 | 507 | 507 | 306 | 306 |
| Male/ | 230/ | 244/ | 232/ | 246/ | 314/ | 332/ | 359/ | 377/ | 223/ | 238/ |
| female | 87 | 73 | 88 | 74 | 132 | 114 | 148 | 130 | 83 | 68 |
| Age (mean±sd) | 56.34 ± 9.42 | 56.34 ± 9.42 | 56.37 ± 9.41 | 56.37 ± 9.41 | 56 ± 9.31 | 56 ± 9.31 | 55.71 ± 9.22 | 55.71 ± 9.22 | 56.45 ± 9.37 | 56.45 ± 9.37 |
| UPDRS | 13.36 ± |  | 13.53 ± |  | 13.42 ± |  | 13.56 ± |  | 13.5 ± 7.95 |  |
| mean±sd (n) | 7.94 (166) | - | 7.99 (174) | - | 7.63 (275) | - | 7.62 (296) | - | (165) | - |
| UPDRS I | 5.77 ± 3.71 | - | 5.84 ± 3.71 | - | 5.56 ± 3.52 | - | 5.81 ± 3.54 | - | 5.86 ± 3.65 | - |
| mean±sd (n) | (172) | - | (179) | - | (284) | - | (304) | - | (172) | - |
| UPDRS II | 7.65 ± 5.4 | - | 7.77 ± 5.44 | - | 7.95 ± 5.41 | - | 7.85 ± 5.34 | - | 7.75 ± 5.38 | - |
| mean±sd (n) | (166) | - | (174) | - | (278) | - | (301) | - | (165) | - |

### B. PRE-PROCESSING

The tri-axial accelerometer integrated in the smartphone records acceleration in the 3 axes (vertical, mediolateral and anteroposterior) during the gait and balance tasks. A 4^th^ order 20 Hz cut-off low-pass Butterworth filter was applied to the 3 accelerometer signals. An additional 3^rd^ order 0.3 Hz cut-off high-pass Butterworth filter was applied to minimize the acceleration variability due to respiration [37]. Signals were then standardized to eliminate the gravity component while maintaining the information from outlier data. According to Pittman et al. [24], 30% of the devices were not held in the correct position and therefore, we additionally calculated the average acceleration signal. Several signals were extracted from the gait recordings including the step series, position along the 3 axes calculated by double integration, velocity and acceleration along the path [38] (Fig. 1).

**Fig. 1.**
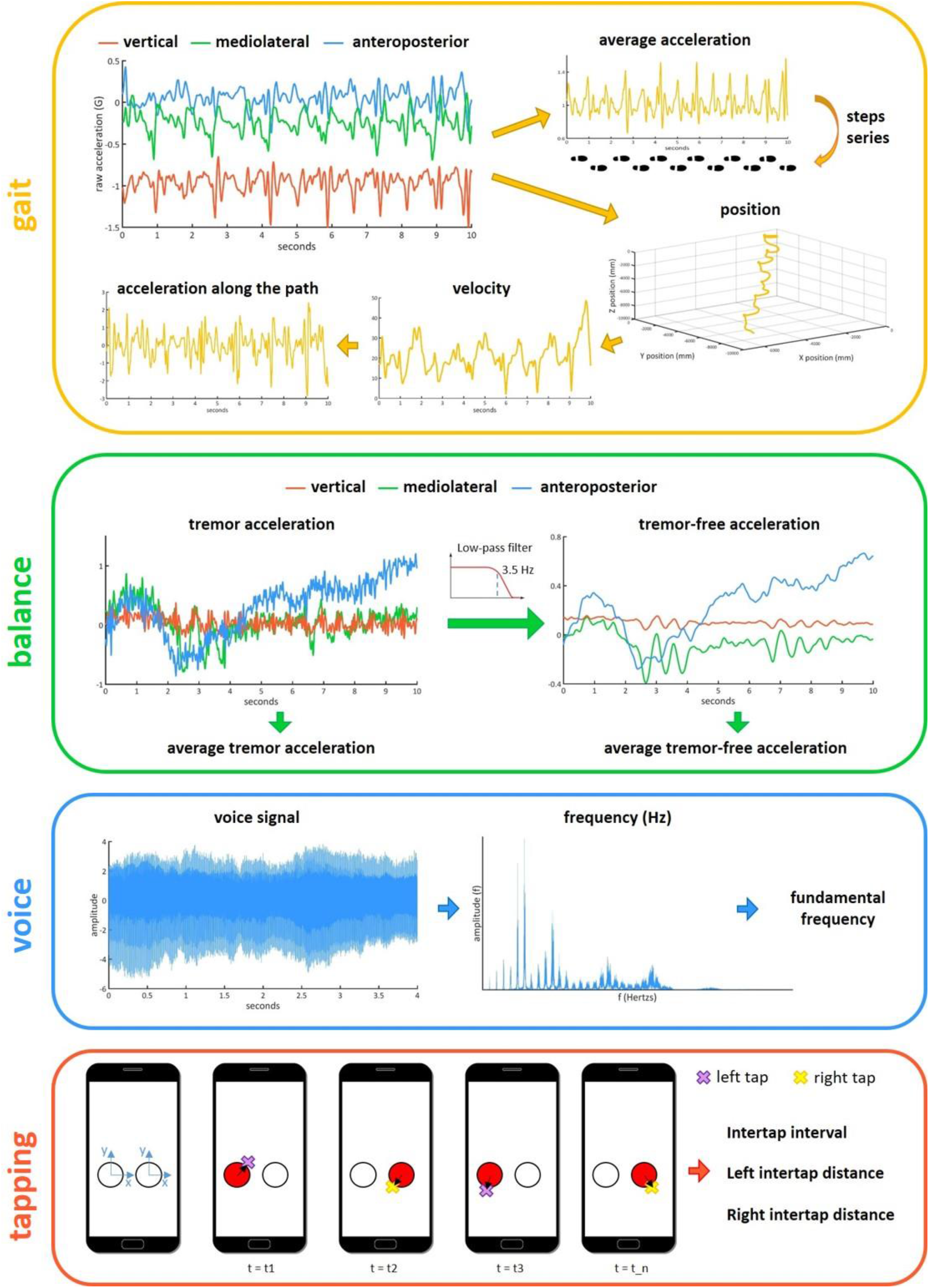
Illustration of signal processing and feature extraction based on the raw data for each task.

Two additional signals were considered for the balance task (Fig. 1). Tremor frequency in PD is estimated to fall in the 4-7 Hz band [39], whereas postural acceleration measures (tremor-free) fall in the 0-3.5 Hz interval. To extract tremor-free measures of postural acceleration, we applied a 3.5 Hz cut-off low-pass Butterworth filter [40].

Voice was recorded at a sample rate of 44.1 Kbps. Pre-processing included a downsampling to 25 KHz and a noise reduction using a 2nd order Butterworth filter with a low-pass frequency at 400 Hz. The fundamental frequency signal was calculated using a Hamming window of 20 ms with 50% overlap, and verified with the software Praat (Fig. 1). Time, frequency and amplitude series were extracted from the voice signals.

Tapping recordings consist of the {x,y} screen pixel coordinates and timestamp for each tap on the screen. Both the inter-tapping interval (time) and the {x,y} inter-tap distance series were computed (Fig. 1). Further details about pre-processing for each task can be found in Appendix I.

### C. FEATURE EXTRACTION

A comprehensive search was conducted in PubMed (https://pubmed.ncbi.nlm.nih.gov/) with the following search terms ((Parkinson’s disease) AND (walking OR gait OR balance OR voice OR tapping) AND (wearables OR smartphones)) to identify features commonly applied for each task and corresponding signals generated. Based on the results of this search, 423, 183, 124 and 43 features were identified and computed using Matlab R2017a from gait [41]–[44], balance [7], [37], [40], [45], voice [25], [26], [46] and tapping data [15], [33], [47], respectively (Table 4-Table 7).

### D. MACHINE LEARNING ALGORITHMS

As a different ML algorithm may provide the best performance for a given task, we evaluated four commonly applied algorithms for differentiation between PD and HC:

1. Least Absolute Shrinkage and Selection Operator (LASSO) is a linear method commonly used to deal with high-dimensional data. LASSO applies a regularization process, where it penalizes the coefficients of the regression variables shrinking some of them to zero. During the feature selection process, those variables with non-zero coefficients are selected to be part of the model [48]. LASSO performs well when dealing with linearly separable data and avoiding overfitting.
2. Random Forest (RF) uses an ensemble of decision trees, where each individual tree outputs the classes. The predicted class is decided based on majority vote. Each tree is built based on a bootstrap training set that normally represents two thirds of the total cohort. The left out data is used to get an unbiased estimate of the classification error and get estimates of feature importance. RF runs efficiently in large datasets and deals very well with data with complicated relationships [49].
3. A Support Vector Machine (SVM) with Radial Basis Function (RBF) kernel with Recursive Feature Elimination (SVM-RFE). An SVM is a linear method whose aim is to find the optimal hyperplane that separates between classes. When data is linearly non-separable, it may be transformed to a higher dimensional space using a non-linear transformation function that spreads the data apart such that a linear hyperplane can be found in that space. Here, we used a radial basis kernel function. RFE is a feature selection method that ranks features according to importance, improving both efficiency and accuracy of the classification model. This model is known to remove effectively non-relevant features and achieve high classification performance [50].
4. Relevance Vector Machine (RVM), which follows the same principles of SVM but provides probabilistic classification. The Bayesian formulation prevents from tuning the hyper-parameters of the SVM. Nonetheless, RVMs use an expectation maximization (EM)-like learning that can lead to local minima unlike the standard sequential optimization (SMO)-based algorithms used by SVMs, that guarantee to find a global optima [51].

### E. FRAMEWORK

The following six experiments were performed to address the questions on the impact of age, sex and comorbidities that may influence task performance on the classification accuracy for each task and on the combination of all tasks for differentiation between PD and HC (Table 2):

**Table 2.** List of experiments indicating their corresponding processing steps.

|  | **Exclude comorbidities** | **Age & sex matching** | **Control for age** | **Control for sex** | **Control for age & sex** |
| --- | --- | --- | --- | --- | --- |
| E1 | - | - | - | - | - |
| E2 | - | X | - | - | - |
| E3 | X | X | - | - | - |
| E4 | X | X | X | - | - |
| E5 | X | X | - | X | - |
| E6 | X | X | X | X | X |

E: Experiment

1. Experiment 1 (E1: all) includes all subjects only restricting the age range (35-75 years old).
2. Experiment 2 (E2: matched) includes subjects after an age and sex matching between PD and HC, where we strictly match one HC for each PD subject with the same age and where possible with the same sex.
3. Experiment 3 (E3: no comorbidities, matched) excludes all comorbidities that may affect task performance (see Appendix I) and strictly matches for age and where possible sex on the remaining subjects.
4. Experiments 4-6 (E4-6): Three additional experiments assess if controlling for age and sex impacts the results. These experiments exclude comorbidities, match for age and sex and control for age and/or sex applying multiple regression to regress out their effects prior to classification: Experiment 4 (E4): no comorbidities, matched, controlled for age; Experiment 5 (E5): no comorbidities, matched, controlled for sex; Experiment 6 (E6): no comorbidities, matched, controlled for age and sex.

As the performance obtained after removing comorbidities and matching for age and sex (E3) provides a relatively unbiased estimate for differentiation between PD and HC, these results were used for selection of the best performing ML algorithm for each task and interpretation of the main outcomes throughout this work. Demographic and clinical information for each experiment are provided in Table 1.

Additionally, to compare the performance of our analyses to those in the literature, we performed an analysis including all data without restricting age range (Table 15) and an analysis including all data and both age and sex as features.

### F. MODEL PERFORMANCE

Data leakage occurs when information of the holdout test set leaks into the dataset used to build the model, leading to incorrect or overoptimistic predictions. Therefore, in every experiment and task, data was initially split into 2/3 of data to build the predictive model and 1/3 of holdout data to validate this model. To build the model, we performed 1000 repetitions of 10-fold cross-validation (CV) in the 2/3 of the data for each classifier to avoid data leakage and increase robustness. The parameter Lambda of the LASSO model was set to 1 and the number of trees for RF to 100. A nested cross-validation was implemented to tune the parameters of the SVM-RFE classifier, following a grid search for the regularization constant (C) ranging from 2^-7^ to 2^7^ and for gamma (γ) ranging from 2^-4^ to 2^4^ for the SVM. For each model, we report the following measures of predictive performance: balanced accuracy (BA), sensitivity, specificity, positive (PPV) and negative predictive value (NPV), mean receiver operating characteristic (ROC) curves with 95% confidence intervals and area under the curve (AUC). Comparisons between models are based on BA.

Once the best predictive model with the highest cross-validation BA was identified using the CV dataset, it was validated using the holdout dataset, reporting the aforementioned performance metrics. In addition, to test whether the BA of the predictive model is higher than chance level (0.5 for binary classification), we ran 1000 permutations randomly permuting the predicted classes, reporting BA at 95% confidence intervals.

## III. RESULTS

### A. CLASSIFIER SELECTION AND RESULTS FOR THE CV DATASET

Four different classifiers (random forest: RF, Least Absolute Shrinkage and Selection Operator: LASSO, support vector machine: SVM, relevance vector machine: RVM-RFE) were applied to each of the four tasks and their combination during the main experiment (E3: no comorbidities, matched for age and sex). Table 9 provides detailed information on the classification performance for each ML algorithm and each task. The ROC curves and corresponding AUC values for the four classifiers for each of the tasks during the cross-validation (CV) step are displayed in Fig. 2A. RF, RVM and SVM-RFE performed similarly across all tasks, whereas LASSO was the classifier performing the poorest. Best performance was achieved on the combination of all tasks using RF (balanced accuracy (BA)): 69.6%), followed by tapping using RVM (BA: 67.9%), balance using RF (BA: 60%), voice using RVM (BA: 56.7%) and gait using SVM-RFE (BA: 56.5%).

**Fig. 2.**
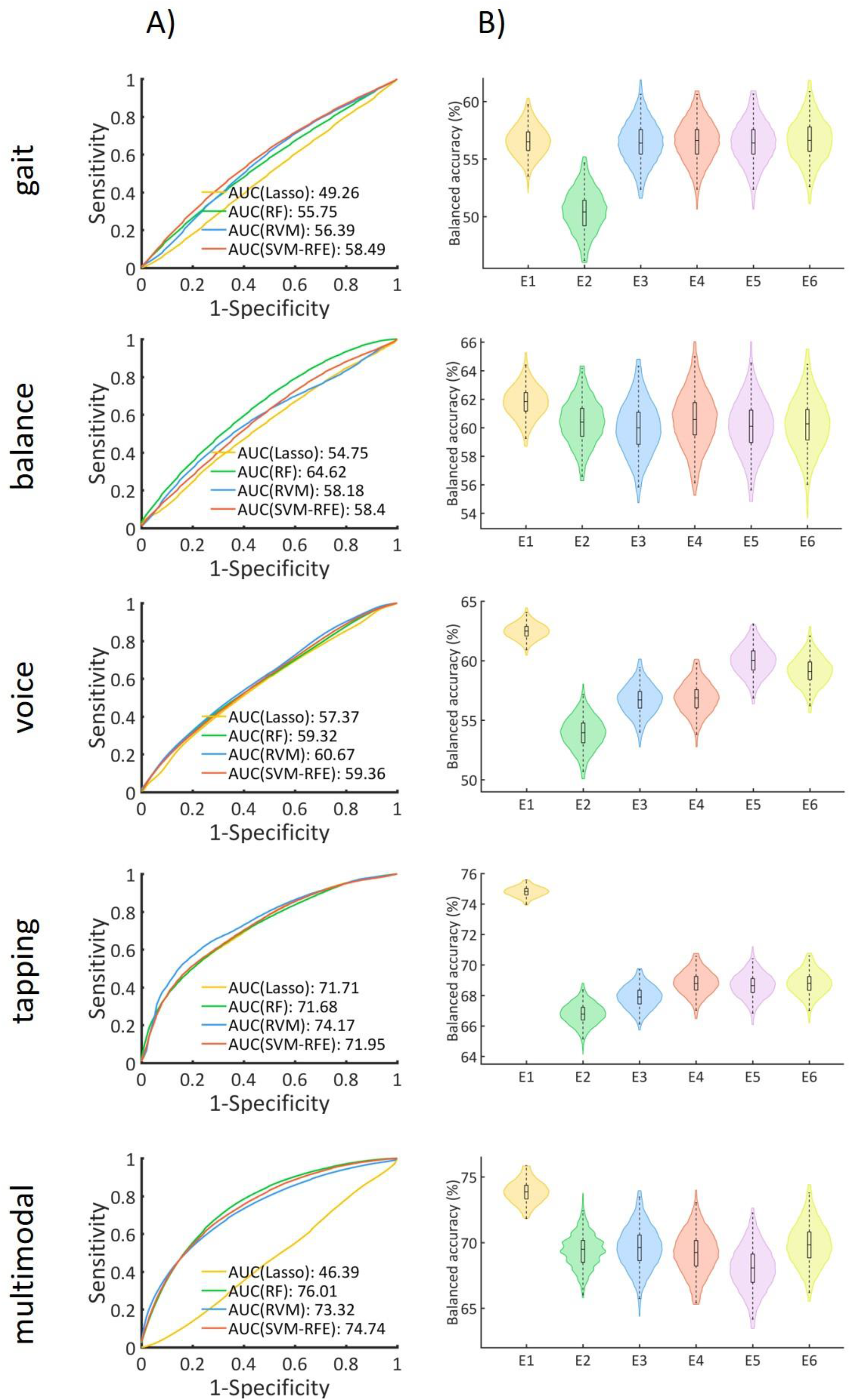
A) ROC curves and AUC values for 4 different classifiers for each task, during the main experiment (E3: no comorbidities, matched). B) Balanced accuracy distributions for each task and experiment (E1-E6). E1: all data. E2: age and sex matched. E3: no comorbidities, age and sex matched. E4: no comorbidities, age and sex matched, controlled for age. E5: no comorbidities, age and sex matched, controlled for sex. E6: no comorbidities, age and sex matched, controlled for age and sex.

### B. COMPARISON OF EXPERIMENTS IN THE CROSS-VALIDATION SETTING

ML algorithms performing best for each task in the main experiment (E3: no comorbidities, matched for age and sex) were applied to corresponding task data of the other five experiments (E1: all subjects, E2: matched for age and sex, E4-6: same as E3 but additionally regressing out the effects of age and/or sex). Classification performance for each task and experiment during the CV and over holdout sets is summarized in Table 3 and Table 10-Table 14. BA distributions for each experiment and task during the CV are displayed in Fig. 2B.

**Table 3.**
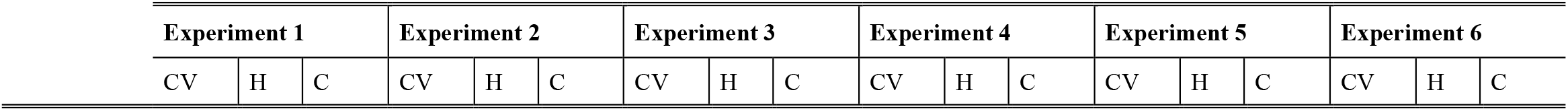
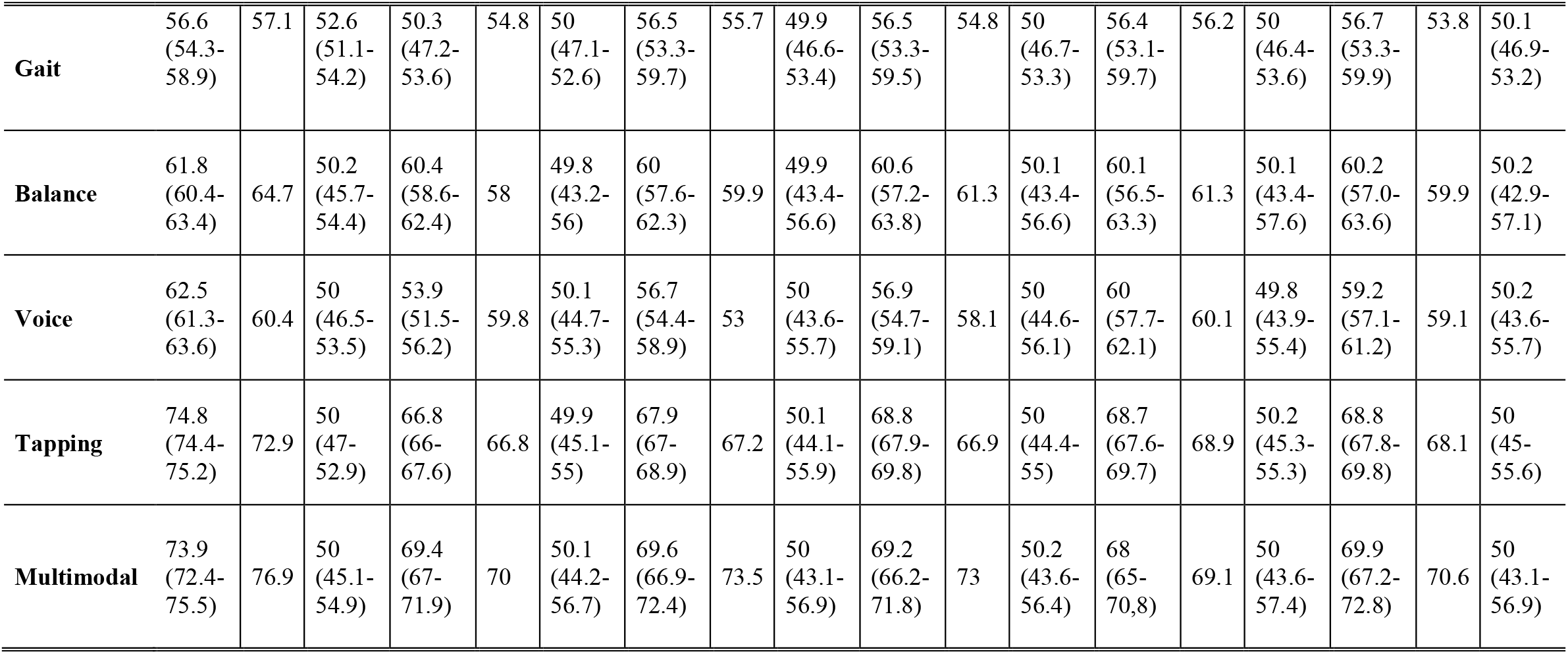
Balanced accuracy results for CV and holdout datasets and chance level at 95%.

| **Experiment 1** |  |  | **Experiment 2** |  |  | **Experiment 3** |  |  | **Experiment 4** |  |  | **Experiment 5** |  |  | **Experiment 6** |  |  |
| --- | --- | --- | --- | --- | --- | --- | --- | --- | --- | --- | --- | --- | --- | --- | --- | --- | --- |
| CV | H | C | CV | H | C | CV | H | C | CV | H | C | CV | H | C | CV | H | C |

|  |  |  |  |  |  |  |  |  |  |  |  |  |  |  |  |  |  |  |
| --- | --- | --- | --- | --- | --- | --- | --- | --- | --- | --- | --- | --- | --- | --- | --- | --- | --- | --- |
| **Gait** | 56.6<br>(54.3-58.9) | 57.1 | 52.6<br>(51.1-54.2) | 50.3<br>(47.2-53.6) | 54.8 | 50<br>(47.1-52.6) | 56.5<br>(53.3-59.7) | 55.7 | 49.9<br>(46.6-53.4) | 56.5<br>(53.3-59.5) | 54.8 | 50<br>(46.7-53.3) | 56.4<br>(53.1-59.7) | 56.2 | 50<br>(46.4-53.6) | 56.7<br>(53.3-59.9) | 53.8 | 50.1<br>(46.9-53.2) |
| **Balance** | 61.8<br>(60.4-63.4) | 64.7 | 50.2<br>(45.7-54.4) | 60.4<br>(58.6-62.4) | 58 | 49.8<br>(43.2-56) | 60<br>(57.6-62.3) | 59.9 | 49.9<br>(43.4-56.6) | 60.6<br>(57.2-63.8) | 61.3 | 50.1<br>(43.4-56.6) | 60.1<br>(56.5-63.3) | 61.3 | 50.1<br>(43.4-57.6) | 60.2<br>(57.0-63.6) | 59.9 | 50.2<br>(42.9-57.1) |
| **Voice** | 62.5<br>(61.3-63.6) | 60.4 | 50<br>(46.5-53.5) | 53.9<br>(51.5-56.2) | 59.8 | 50.1<br>(44.7-55.3) | 56.7<br>(54.4-58.9) | 53 | 50<br>(43.6-55.7) | 56.9<br>(54.7-59.1) | 58.1 | 50<br>(44.6-56.1) | 60<br>(57.7-62.1) | 60.1 | 49.8<br>(43.9-55.4) | 59.2<br>(57.1-61.2) | 59.1 | 50.2<br>(43.6-55.7) |
| **Tapping** | 74.8<br>(74.4-75.2) | 72.9 | 50<br>(47-52.9) | 66.8<br>(66-67.6) | 66.8 | 49.9<br>(45.1-55) | 67.9<br>(67-68.9) | 67.2 | 50.1<br>(44.1-55.9) | 68.8<br>(67.9-69.8) | 66.9 | 50<br>(44.4-55) | 68.7<br>(67.6-69.7) | 68.9 | 50.2<br>(45.3-55.3) | 68.8<br>(67.8-69.8) | 68.1 | 50<br>(45-55.6) |
| **Multimodal** | 73.9<br>(72.4-75.5) | 76.9 | 50<br>(45.1-54.9) | 69.4<br>(67-71.9) | 70 | 50.1<br>(44.2-56.7) | 69.6<br>(66.9-72.4) | 73.5 | 50<br>(43.1-56.9) | 69.2<br>(66.2-71.8) | 73 | 50.2<br>(43.6-56.4) | 68<br>(65-70.8) | 69.1 | 50<br>(43.6-57.4) | 69.9<br>(67.2-72.8) | 70.6 | 50<br>(43.1-56.9) |

**Table 4.**
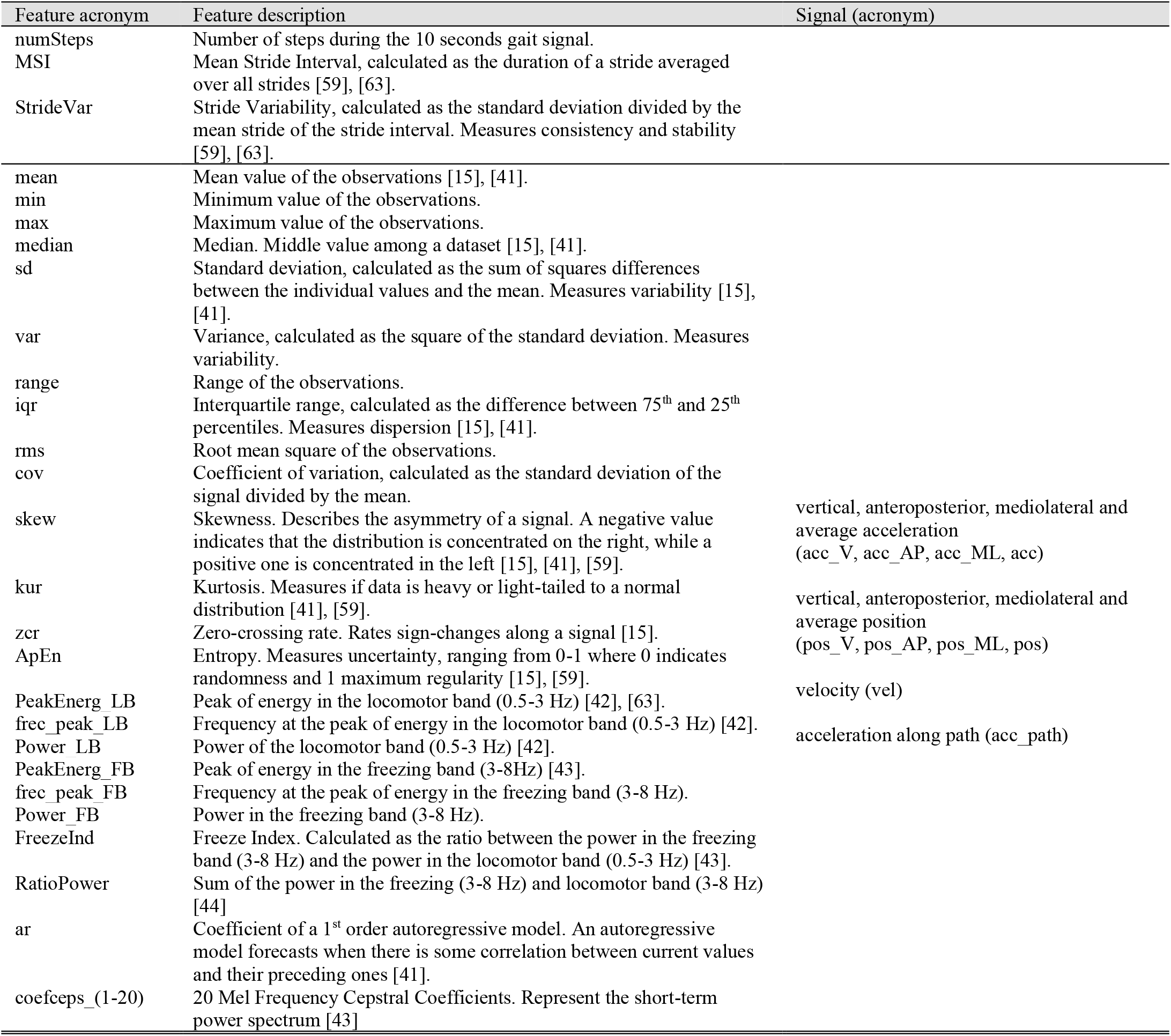
List of gait features.

In the CV, E1 (all data) resulted in the highest but modest BA for all tasks (gait: 56.6%; balance: 61.8%; voice: 62.5%; tapping: 74.8; multimodal combining all four tasks: 73.9%). Removal of comorbidities in E3 had a marginal effect on BA as compared to E2 (matched for age and sex) with increased BA for gait (E2: 50.3%; E3: 56.5%), voice (E2: 53.9%; E3: 56.7%) and tapping (E2: 66.8%; E3: 67.9%) but lower BA for balance (E2: 60.4%; E3: 60.0%). After additionally regressing out the effects of age and/or sex (E4-E6) the change in the BA was negligible for all tasks (< 1%) except for voice when regressing out sex (E3: 56.7%; E5: 60%) and both age and sex (E3: 56.7%; E6: 59.2%) (Table 3, Table S7-S11).

Analyses including all data without trimming for age range led to the highest accuracy of 74.4% using tapping data, followed by 72.7% for the multimodal case and 58%, 52.9% and 51% for balance, voice and gait data respectively. In all cases specificity was close to 100% whereas sensitivity was exceedingly low (Table 16-Table 20). When including both age and sex as additional features, accuracy increased to 80.8% for tapping data, 75.3% for the multimodal case and 73.1%, 69% and 57% for voice, balance and gait data respectively with high specificities and low sensitivities.

### C. RESULTS FOR THE HOLDOUT DATASET

Best performing classifiers trained on the 2/3 of the initial dataset used for cross-validation were applied to the 1/3 holdout dataset. Results for the holdout dataset were highly similar to the CV results (Table 3, Table S7-S11). All results are summarized in Fig. 3 and Table 3. The multimodal combination of all tasks resulted in the best performance for differentiation of PD and HC in the holdout cohort (BA: 73.5%) followed by the tapping features (67.2%). Voice features achieved the lowest BA of 53% followed by gait (55.7%) and balance (59.9%) features (Table 3). For the base experiment E3, the difference in BA between CV and holdout sets was less than 1% for all tasks except for a 3.7% reduction in BA for voice data and a 3.9% increase for the multimodal feature combination. Exclusion of comorbidities resulted in only minor changes for gait, balance and tapping (<2%) with a 6.8% drop only observed using voice data and a 3.5% increase for the multimodal case. BA performance for all tasks increased by 1.4% (gait) to 7.4% (voice) for all tasks when using the dataset only restricting the age range (E1) as compared to E3. No systematic effects of additionally controlling for age and/or sex prior to classification (E4-E6) were observed with BA changes being small and inconsistent across tasks and experiments.

**Fig. 3.**
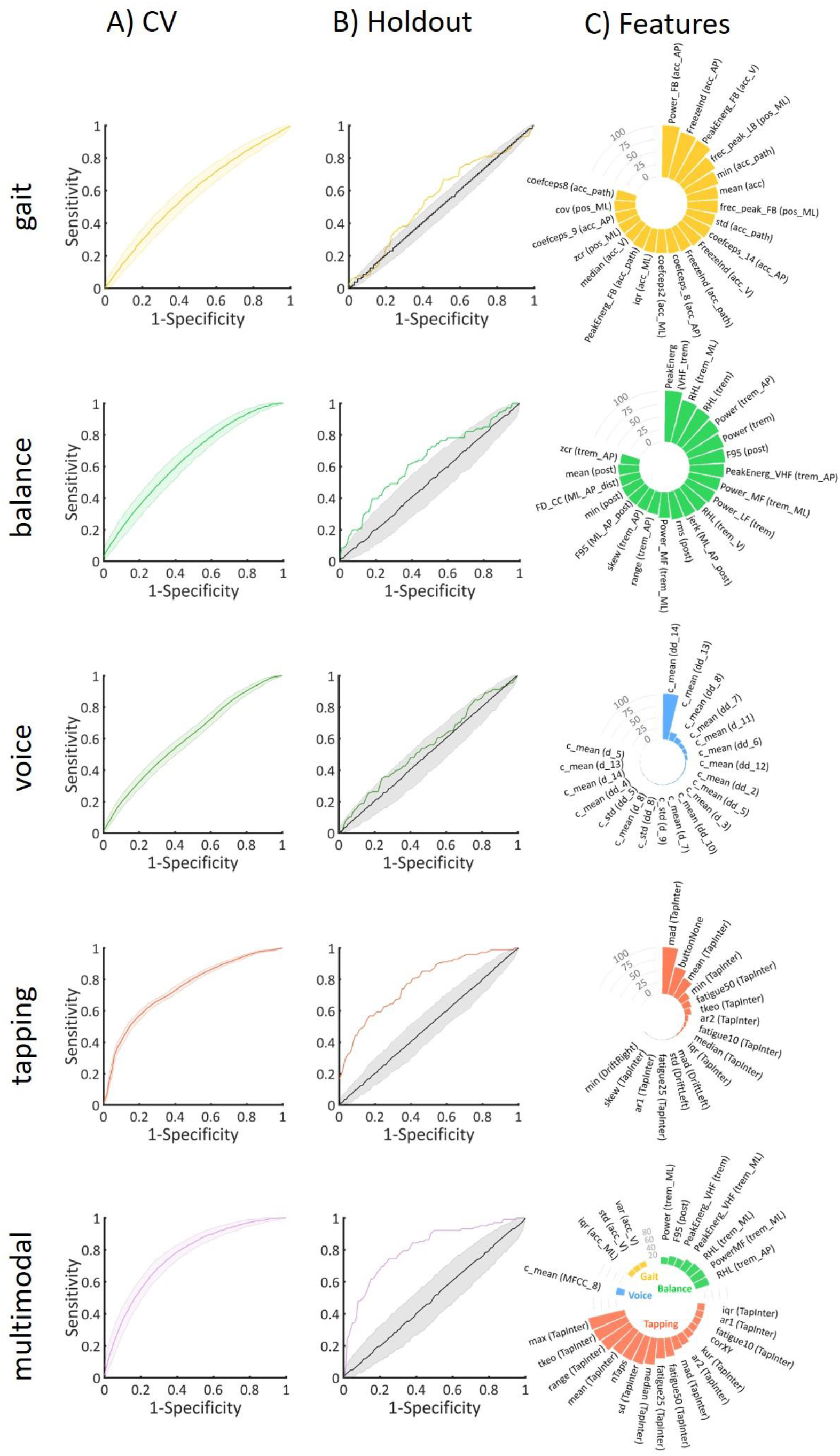
A) ROC curves at 95% CI during CV. B) ROC curves at 95% CI during validation of holdout set and at the chance level. C) Scaled average weights of features for each task for the main experiment (E3: no comorbidities, matched). Gait) acc - average acceleration, acc_path – acceleration along path, AP – anteroposterior, FB – freezing band, LB – locomotor band, ML – mediolateral, pos – position, V – vertical, vel – velocity. Balance) trem – tremor, post – postural, dist – distance, LF – low frequency, MF – medium frequency, VHF – very high frequency, RHL – ratio between high and low frequency, F95 –frequency containing 95% of the power spectrum. Voice) c – cepstral coefficient, d – 1^st^ derivative of cepstral coefficient, dd – 2^nd^ derivative of cepstral coefficient. Tapping) TapInter – tap interval. For details on features refer to Appendix I.

Analyses including all data without trimming for age range reached the highest accuracy in the holdout set of 73.3% using multimodal features, followed by 71.1% for the tapping task and 55.8%, 52.6% and 51.6% for balance, voice and gait data respectively (Table 16-Table 20). When including both age and sex as additional features, accuracy in the holdout data raised to 78.9% for tapping data, 75.9% for the multimodal case and 74.6%, 66% and 58.3% for voice, balance and gait data respectively with very high specificities and very low sensitivities.

### D. PREDICTIVE FEATURES

Best performance during CV for the main experiment E3 was achieved using the multimodal set of features. Fig. 3 shows the scaled average absolute feature weights for RVM and SVM-RFE and the scaled average importance scores for RF, calculated with the out-of-bag (OOB) permuted predictor delta error across 1000 repetitions during the CV. Features with the highest importance scores belong to the tapping task followed by the balance task. Tapping features with the highest importance scores comprised the range of intertap interval (100), maximum value of the intertap interval (99.8) and Teager-Kaiser energy operator of the intertap interval (83.2). Balance features with highest importance scores were the power ratio between high (3.5-15 Hz) and low (0.15-3.5 Hz) frequency for anteroposterior acceleration (31.5) and energy in the medium frequency band for mediolateral acceleration (25.3). Gait and voice tasks had the least contributions in terms of importance scores.

## IV. DISCUSSION

Here, we systematically evaluated the ability of four commonly applied DB tasks to differentiate between PD and HC in a self-administered remote setting. Our findings indicate that, depending on the constellation, not accounting for confounds in PD digital biomarker task data may lead to under-but also over-optimistic results.

### A. IDENTIFICATION OF PARKINSON’S DISEASE

Out of the four evaluated machine learning algorithms, similar performance was achieved for all classifiers except LASSO which showed the poorest performance. Whereas some previous studies using the mPower dataset selected different algorithms according to tasks [25], [26], others simply applied a single classifier [28], [30]. No single classifier performed best for all four tasks in our study. This is in line with previous research showing that the selection of the classifier depends mainly on the type and complexity of the data [52], [53]. For instance, RF, RVM and Gaussian SVM are non-linear algorithms, offering more flexibility regarding the type of data. On the contrary, LASSO is a linear classifier and thus, its performance depends on whether the data is linearly separable. Whereas the generalizability of this observation is limited by the use of only one linear classifier, it may point to a better usability of non-linear approaches for classification of digital assessments.

For discrimination of PD and HC, combination of all tasks reached a BA of 74%, followed by tapping that achieved 67%, outperforming other tasks which were close to chance level. These results are in line with previous literature using the mPower dataset, where tapping reached the highest accuracies and gait and voice were closer to chance level [30]. Several studies reported higher accuracies for this type of data [24], [28]. Yet, these studies followed certain “optimistic” approaches as discussed below.

### B. POTENTIAL CONFOUNDERS

Exclusion of comorbidities resulted in increased accuracies by a few percent, suggesting that other diseases may add more variability to the signal. Prediction performances considerably decreased for all tasks after matching for age and sex indicating the importance of controlling for such confounds in DB data. When including all data without trimming age range, accuracies greatly increase. Nonetheless, specificity values are exceedingly high whereas sensitivity values are vastly low. This indicates a greater prediction ability for the HC group, which is considerably larger than the PD group for subjects under 35 years old. Including age and sex as part of the features resulted in further accuracy increases, yet with very low sensitivities. Since the dataset is strongly slanted toward young HC, the model is most likely distinguishing HC based on age and gender in this case. Such effects may also explain the high accuracies in some of the previous studies using mPower dataset, where no proper matching for these confounds was performed, age and/or sex were used as features despite a large imbalance across groups or non-balanced accuracies were reported [24], [26], [28], [35]. In example, in the overall mPower dataset HC outnumber PD by a factor of five and age and sex alone provide a high discrimination accuracy between PD and HC with PD being on average 28 years older and more often female (34% of PD vs 19% of HC). Our findings are also in line with previous studies demonstrating a similarly strong decrease in accuracies when accounting for respective confounds. Neto et al. [54] studied the effect of confounders on gait data. They reached very high accuracy when not accounting for confounders, compared with a very modest accuracy when using unconfounded measures. Schwab and Karlent [25] performed analysis with all the tasks from the mPower dataset with and without including age and sex, the latter resulting in a similarly low accuracy as in our study.

For all classification experiments, we used only one recording per subject to prevent the classifier from detecting the idiosyncrasies of each subject rather than specific PD related symptoms [30]–[32]. Single measures are likely to contain more noise due to higher variation in task administration as well as in individual performance in a poorly-controlled setting [55]. Using multiple time points may therefore further increase the discrimination between PD and HC as demonstrated in several previous studies [30]– [32]. Yet, our results in this respect highlight the need of further understanding and better control of the individual parameters which impact the task performance during a single administration.

### C. PREDICTORS OF PARKINSON’S DISEASE

Features with largest weights in the multimodal discrimination between PD and HC were derived from the tapping task. These features mostly related to the inter-tapping interval (time), presumably reflecting bradykinesia-like symptoms. These results are in line with previous studies, where tapping features related to speed and accuracy had the strongest correlation with clinical scores [56], [57]. Balance task features related to tremor measures had larger weights than postural ones. In addition, features from the frequency domain had greater weights than spatiotemporal features. Spatiotemporal features have been extensively studied and applied, due to their ease of computation and interpretability [58]. However, these features offer information limited primarily to leg movement, whilst frequency features add information regarding asymmetry and variability. Furthermore, balance features with higher weights belonged to the mediolateral and anteroposterior signals, related to stability. Even though gait had limited contribution to the classification accuracy, acceleration features had the highest weights from this task. This observation is in line with previous findings where acceleration proved to better capture PD-related gait changes [59]. In line with some previous studies, features with the highest weights from the voice task were all based on Mel Frequency Cepstral Coefficients which can detect subtle changes in speech articulation that are common in PD [60], [61].

### D. LIMITATIONS AND FURTHER RESEARCH

Whereas sensors-integrated in smartphones open new opportunities for at-home continuous, reliable, non-invasive and low-cost monitoring of PD, our finding highlight the need for further development, optimization and standardization of specific measures for such applications.

The interpretation of our findings is limited by several aspects, including the lack of standardization, poor control of environmental and medication effects during performance of the tasks and intentionally or unintentionally incorrect information provided by the participants. In addition, removal of comorbidities and matching for age and sex led to exclusion of about 50% of data, which may affect the training of classifiers [54].

## Data Availability

The m-Power dataset used for this article is available upon registration from Synapse at: https://www.synapse.org/#!Synapse:syn4993293/

## APPENDIX I

### SUPPLEMENTARY METHODS

#### A. DATA CLEANING

MPower dataset offers demographic, PDQ8 and MDS-UPDRS surveys and task-based data. The demographics table contains data for 6805 subjects. In order to establish a diagnosis, participants had to select “true” or “false” to the following question “Have you been diagnosed by a medical professional with Parkinson Diseaseã”. According to this answer, they are classified as Parkinson’s Disease (PD) or Healthy Control (HC). Some subjects left this question unanswered and thus they were discarded from further analysis. Those subjects classified as PD which did not completed the PDQ8 and MDS-UPDRS questionnaire were also excluded. Subjects with no information on age, sex or any task data were also removed, resulting in 6614 subjects. Those empty, null or corrupted files for each task were deleted, resulting in 2807 subjects with gait and balance data, 4925 with voice data and 6366 with tapping data. Since a large number of subjects are HC under 35 years old, our analysis focused on a subset of subjects within the age range of 35 to 75 years old, leading to 1435 subjects with gait and balance data, 2186 subjects with voice data and 2644 subjects with tapping data. Finally, all subjects with inconsistencies for each of the tasks were discarded (i.e., subjects that reported not to have been diagnosed with Parkinson’s disease but filled in PD medication questions, year of diagnosis of PD, surgery or deep brain stimulation). This last elimination resulted in 1416 subjects with gait and balance data, 2153 subjects with voice data and 2600 subjects with tapping data.

#### B. SIGNALS LENGTH

Gait task consists of walking 20 steps in a straight line. In order to analyse the same signal length for each subject, we examined how many subjects had gait data for different time durations. We observed that after 10 seconds, participation was dropping heavily. Therefore, we selected a time length of 10 seconds and discarded those participants with shorter signals. Following the same reasoning, we chose voice signals of 7 seconds, trimming the first second and last two seconds, and tapping signals of 20 seconds. Similarly, balance task consists of standing still for 30 seconds although just 20 seconds were selected. Nonetheless, whereas gait, voice and tapping are independent tasks, and therefore they are started by the user, balance task starts straight after the gait task. This is, as soon as the gait task ends, the app plays out loud “turn around and stand still for 30 seconds”. As a result, most of the balance recordings include initial slots of noise, which most likely coincide with the time that subjects listen to the instructions, react, turn around and start the task. Therefore, we trimmed the first 5 seconds of the signal, resulting in balance signals of 15 seconds for all subjects. Final number of subjects consisted of 1397 subjects with gait data, 1415 subjects with balance data, 2150 subjects with voice data and 2600 subjects with tapping data.

#### C. PRE-PROCESSING AND SIGNAL EXTRACTION

Gait and balance data consists on vertical (V), anteroposterior (AP) and mediolateral (ML) acceleration signals. For these 3 gait acceleration signals, we applied a Butterworth low pass filter with cut-off frequency at 20 Hz followed by a 3° order high pass filter at 0.3 Hz. According to Pittman et al. [24], around 30% of devices were not held in the correct position. Therefore, the greatest gravitational displacement is not always along the vertical axis. Then, we standardized these three signals and calculated an additional average acceleration signal. Based on the standardized acceleration signal, we extracted the step series. We calculated position signals along the three axes by double integrating the acceleration signals and the average position. Then, we extracted velocity and acceleration along the path by derivation [38].

Balance acceleration signals were filtered with a low pass Butterworth filter at 20 Hz. Since tremor in PD usually falls in the 4-7Hz frequency band [39], [40], the interval 0-3.5 Hz is considered for tremor-free or postural acceleration measures. Hence, we applied a Butterworth filter at 3.5 Hz to extract postural acceleration measures. We also calculated the average of the tremor acceleration in the 3 axes and the average of the postural acceleration in the 3 axes.

Voice signals were recorded at a sample frequency of 44.1 KHz. We downsampled the signal to 25KHz, applied a second order Butterworth filter with cut-off frequency at 400 Hz followed by a pre-emphasis FIR filter for noise reduction and correct for distortions. We extracted the fundamental frequency (f0) series, which was verified with the Praat software.

Tapping recordings consists of the {x,y} screen pixel coordinates and timestamp for each tap on the screen. Signals derived out of these recordings were the inter-tapping interval (time) and the {x,y} inter-tap distance series.

#### D. FEATURE EXTRACTION

1. GAIT We extracted 11 signals from the original accelerometer recordings during gait tasks. These are V, AP and ML acceleration, the step series, the average of the acceleration in the three axes, the V, AP and ML position, the average position in the three axes, the velocity and the acceleration along the path. Table 4 collects a list of features extracted for these signals along with their acronyms.
2. BALANCE Balance signals consist in the V, AP and ML tremor acceleration (4-7 Hz), the average of these 3 signals, the V, AP and ML postural acceleration (0-3.5 Hz) and the average of these 3 signals. We extracted displacement-related postural features from ML, AP and average of both distance signals, following the formulation in Martinez-Mendez et al. [37] (Table 5).

**Table 5.** List of balance features.
3. VOICE Most of voice features were extracted following the formulation in Tsanas et al. [46]. Tsanas et al. state that the period (T) signal provides different information than f0. Therefore, we additionally extracted the T series. Further signals include glottis quotient and 14 Mel Frequency Cepstral Coefficients (MFCCs), including the 0^th^ coefficient and the log-energy of the signal, along with their associated delta and delta-delta coefficients as applied in the Voicebox Matlab ToolBox [62] (Table 6).

| Acronym | Description | Signal (acronym) |
| --- | --- | --- |
| mean | Mean value of the observations. |  |
| min | Minimum value of the observations. |  |
| max | Maximum value of the observations. |  |
| median | Median value of the observations. |  |
| sd | Standard deviation, calculated as the sum of squares differences between the individual values and the mean. Measures variability. |  |
| var | Variance, calculated as the square of the standard deviation. Measures variability. |  |
| range | Range of the observations. | vertical, anteroposterior, mediolateral and average tremor acceleration (trem_V, trem_AP, trem_ML, trem) |
| iqr | Interquartile range, calculated as the difference between 75<sup>th</sup> and 25<sup>th</sup> percentiles. Measures dispersion. |  |
| rms | Root mean square of the observations. |  |
| cov | Coefficient of variation, calculated as the standard deviation of the signal divided by the mean. | vertical, anteroposterior, mediolateral and average postural acceleration (post_V, post_AP, post_ML, post) |
| skew | Skewness. Describes the asymmetry of a signal. A negative value indicates that the distribution is concentrated on the right, while a positive one is concentrated in the left. |  |
| kur | Kurtosis. Measures if data is heavy or light-tailed to a normal distribution. |  |
| zcr | Zero-crossing rate. Rates sign-changes along a signal. |  |
| ApEn | Entropy. Measures uncertainty, ranging from 0-1 where 0 indicates randomness and 1 maximum regularity. |  |
| Power_MF | Power of the medium frequency band (4-7Hz) [40]. |  |
| PeakEnerg_VHF | Peak of energy in the very high frequency band (>7Hz) |  |
| frec_peak_HF | Frequency at the peak of energy in the high frequency band (>4Hz) [40]. | vertical, anteroposterior, mediolateral and average tremor acceleration (trem_V, trem_AP, trem_ML, trem) |
| Power | Power between 3.5-15Hz |  |
| Power_LF | Power in the low frequency band (0.15-3.5Hz) |  |
| RHL | Ratio between the power between 3.5-15Hz and power between 0.15-3.5Hz [40]. |  |
| CFREQ | Centroidal frequency for postural measures. Also known as zero-crossing frequency [37], [40], [45]. | anteroposterior, mediolateral and average postural acceleration (post_AP, post_ML, post) |
| FREQD | Frequency of dispersion of the power spectrum for postural measures [37], [40], [45]. | mediolateral-anteroposterior average postural acceleration (ML_AP_post) |
| jerk | Average jerk. Measures vibration as the rate of change in acceleration. Calculated as the derivative of acceleration with respect to time [37], [40]. | vertical, anteroposterior, mediolateral and average postural acceleration (post_V, post_AP, post_ML, post) |
| TotalPower | Energy between 0.15-3.5Hz for postural measures [37]. |  |
| F50 | Frequency containing 50% of the total power for postural measures [37], [40]. | mediolateral-anteroposterior average postural acceleration (ML_AP_post) |
| F95 | Frequency containing 95% of the total power for postural measures [37], [40]. |  |
| MDIST | Represents the average distance from the center to each AP and ML points [40], [45]. |  |
| RDIST | Root Mean Square distance from the mean center [45]. |  |
| TOTEX | Total excursions is the total length of the path. Calculated as the sum of distances between consecutive points [45]. | mediolateral, anteroposterior and average of mediolateral and anteroposterior distance (ML-dist, AP_dist, ML_AP_dist) |
| MVELO | Mean velocity is the average velocity of the center path, calculated as the TOTEX divided by the time [40], [45]. |  |
| MFREQ | The mean frequency is the rotational frequency with a radius equal to the mean distance [37], [45]. |  |
| AREA_CC | The 95% confidence circle area is the area of a circle enclosing all points in the AP-ML plane with 95% confidence [37], [45]. |  |
| AREA_CE | The 95% confidence ellipse area is the area of an ellipse enclosing all points in the AP-ML plane with 95% confidence [37], [40], [45]. |  |
| AREA_SW | Sway area calculated as the area enclosing the acceleration path [37], [40], [45]. |  |
| FD | The fractal dimension indicates the degree to which a curve fills the enclosed metric space [37], [45]. | average of mediolateral and anteroposterior distance (ML_AP_dist) |
| FD_CC | Fractal dimension based on the 95% confidence circle area [37], [45]. |  |
| FD_CE | Fractal dimension based on the 95% confidence ellipse area [37], [45]. |  |

**Table 6.** List of voice features.
4. TAPPING We considered a set of features computed from the inter-tapping interval (time) and the {x,y} inter-tap distance signals, according to Bot et al. [47] (Table 7).

| Feature | Feature description | Signal (acronym) |
| --- | --- | --- |

| acronym |  |  |
| --- | --- | --- |
| amp | Average amplitude [46]. |  |
| shim | Absolute shimmer [15], [26], [46]. |  |
| shdb | Shimmer in logarithmic domain [46]. |  |
| apq3 | 3 point amplitude perturbation quotient in percentage [46]. |  |
| apq5 | 5 point amplitude perturbation quotient in percentage [46]. |  |
| fm | Frequency modulation [46]. |  |
| hnr_mean | Mean of the harmonic to noise ratio, which indicates the amount of noise [15], [26], [46]. |  |
| hnr_std | Standard deviation of the harmonic to noise ratio [46]. |  |
| rpde | Recurrence period density entropy. Characterizes the deviation from signal periodicity [15], [46]. |  |
| DFA | Detrended Fluctuation Analysis, which describes turbulent noise [15], [46]. |  |
| mean | Mean value [15], [46]. | fundamental frequency (f0), amplitude (amp), Teager Kaiser Energy Operator of the fundamental frequency (tkeo_f0), open quotient (oq), glottis quotient open (gqo), glottis quotient closed (gqc) |
| sd | Standard deviation [15], [46]. |  |
| jitt | Absolute jitter [15], [46]. |  |
| jitta | Relative or local jitter [46]. |  |
| rap | Relative average perturbation [46]. | fundamental frequency (f0), period (T) |
| ppq5 | Perturbation quotient using 5 point (cycles) [46]. |  |
| range | Range [46]. |  |
| tkeo_p25 | 25<sup>th</sup> percentile of the Teager-Kaiser Energy Operator [46]. | fundamental frequency (f0) |
| tkeo_p75 | 75<sup>th</sup> percentile of the Teager-Kaiser Energy Operator [46]. |  |
| ApEn | Pitch Period Entropy. Quantifies the impaired control of stable pitch during a sustained phonation [15], [46]. |  |
| p5 | 5<sup>th</sup> percentile [46]. | Teager Kaiser Energy Operator of the fundamental frequency (tkeo_f0), open quotient (oq), glottis quotient open (gqo), glottis quotient closed (gqc) |
| p95 | 95<sup>th</sup> percentile [46]. |  |
| c_mean | Mean of the Mel Frequency Cepstral Coefficients (MFCCs) coefficients, log-energy of the signal and the first and second derivatives of the MFCCs [15], [26], [46]. | log energy (log), 0<sup>th</sup> order cepstral coefficient (0th), 1-12<sup>th</sup> Mel Frequency Cepstral Coefficients (MFCC_(1-12), 1-14<sup>th</sup> deltas (d_(1-14)), 1-14<sup>th</sup> delta-delta (dd_(1-14))) |
| c_std | Standard deviation of the MFCCs coefficients, log-energy of the signal and the first and second derivatives of the MFCCs [26], [46]. |  |

**Table 7.** List of tapping features.

| Feature acronym | Feature description | Signal (acronym) |
| --- | --- | --- |
| nTaps | Number of taps [47], [64]. |  |
| buttonNone | Frequency of tapping outside the button [47], [64]. |  |
| corXY | Correlation between X and Y touchscreen coordinates [47], [64]. |  |
| mean | Mean value of the observations [15], [47], [64]. | Intertap interval (TapInter), Leftdrift (DriftLeft), Right drift (DriftRight) |
| min | Minimum value of the observations [15], [47], [64]. |  |
| max | Maximum value of the observations [15], [47], [64]. |  |
| median | Median value of the observations [15], [47], [64]. |  |
| mad | Median absolute deviation [47], [64]. |  |
| sd | Standard deviation [15], [47], [64]. |  |
| range | Range of the observations [15], [47], [64]. |  |
| iqr | Interquartile range [47], [64]. |  |
| cov | Coefficient of variation [15], [47], [64]. |  |
| skew | Skewness [47], [64]. |  |
| kur | Kurtosis [47], [64]. |  |
| tkeo | Teager-Kaiser Energy Operator. Measures energy variation [15], [47], [64]. |  |
| dfa | Detrended Fluctuation Analysis. Measures changes in the signal [15], [47], [64]. |  |
| ar1 | Coefficient of an autoregressive model at lag 1. Indicates associations between intertap intervals [15], [47], [64]. | Intertap interval (TapInter) |
| ar2 | Coefficient of an autoregressive model at lag 2. Indicates associations between intertap intervals [15], [47], [64]. |  |
| fatigue10 | Increase in the mean intertap interval from the first 10% to the last 10% taps [15], [47], [64]. |  |
| fatigue25 | Increase in the mean intertap interval from the first 25% to the last 25% taps [15], [47], [64]. |  |
| fatigue50 | Increase in the mean intertap interval from the first 50% to the last 50 % taps [15], [47], [64]. |  |

### E. COMORBIDITIES

Comorbidities selected for removal in the experiments E3-E6 include “Alzheimer Disease or Alzheimer dementia”, “Dementia”, “Schizophrenia or Bipolar Disorder”, “Alcoholism”, “Multiple Sclerosis”, “Leukemia or Lymphoma”, “Acute Myocardial Infarction/Heart Attack”, “Stroke/Transient Ischemic Attack”, “Breast Cancer”, “Colorectal Cancer”, “Prostate Cancer”, “Lung Cancer”, “Endometrial/Uterine Cancer”, “Any other kind of cancer OR tumor”, “Heart Failure/Congestive Heart Failure”, “Ischemic Heart Disease”. These comorbidities were removed since they may lead to brain damage or to undertake chemotherapy or other therapy, which might induce brain changes.

### F. MEDICATION STATUS

Table 8 shows the number of subjects that performed the task just before taking their medication, after taking their medication, at another random time, number of those who were not taking any medication and number of those who did not give any information about their medication status.

**Table 8.** Medication status at the time of performing the tasks.

|  |  |  | PD/Total | Before | After | Another | No Med | Empty |
| --- | --- | --- | --- | --- | --- | --- | --- | --- |
| E1 | Gait | All | 653/2711 | 154 | 166 | 259 | 63 | 11 |
|  |  | CV | 436/2711 | 99 | 120 | 169 | 39 | 9 |
|  |  | Holdout | 217/903 | 55 | 46 | 90 | 24 | 2 |
|  | Balance | All | 655/2747 | 156 | 166 | 257 | 64 | 12 |
|  |  | CV | 437/1832 | 103 | 99 | 186 | 41 | 8 |
|  |  | Holdout | 218/915 | 53 | 67 | 71 | 23 | 4 |
|  | Voice | All | 965/4799 | 222 | 229 | 396 | 94 | 24 |
|  |  | CV | 644/3200 | 140 | 159 | 265 | 62 | 18 |
|  |  | Holdout | 321/1599 | 82 | 70 | 131 | 32 | 6 |
|  | Tapping | All | 1054/6221 | 237 | 237 | 446 | 106 | 28 |
|  |  | CV | 703/4148 | 156 | 157 | 299 | 72 | 19 |
|  |  | Holdout | 351/2073 | 81 | 80 | 147 | 34 | 9 |
| E2 | Gait | All | 373/746 | 91 | 101 | 135 | 39 | 7 |
|  |  | CV | 249/498 | 62 | 70 | 86 | 28 | 3 |
|  |  | Holdout | 124/248 | 29 | 31 | 49 | 11 | 4 |
|  | Balance | All | 376/752 | 99 | 96 | 139 | 36 | 6 |
|  |  | CV | 251/502 | 68 | 60 | 97 | 21 | 5 |
|  |  | Holdout | 125/250 | 31 | 36 | 42 | 15 | 1 |
|  | Voice | All | 534/1068 | 135 | 131 | 205 | 48 | 15 |
|  |  | CV | 356/712 | 94 | 89 | 127 | 34 | 12 |
|  |  | Holdout | 178/356 | 41 | 42 | 78 | 14 | 3 |
|  | Tapping | All | 608/1216 | 134 | 135 | 262 | 59 | 18 |
|  |  | CV | 406/812 | 83 | 92 | 182 | 39 | 10 |
|  |  | Holdout | 202/404 | 51 | 43 | 80 | 20 | 8 |
| E3 | Gait | All | 317/634 | 82 | 84 | 117 | 28 | 6 |
|  |  | CV | 212/424 | 54 | 53 | 82 | 19 | 4 |
|  |  | Holdout | 105/210 | 28 | 31 | 35 | 9 | 2 |
|  | Balance | All | 320/640 | 76 | 89 | 116 | 32 | 7 |
|  |  | CV | 214/428 | 48 | 53 | 87 | 22 | 4 |
|  |  | Holdout | 106/212 | 28 | 36 | 29 | 10 | 3 |
|  | Voice | All | 446/892 | 112 | 103 | 190 | 34 | 7 |
|  |  | CV | 298/596 | 75 | 71 | 125 | 21 | 6 |
|  |  | Holdout | 148/296 | 37 | 32 | 65 | 13 | 9 |
|  | Tapping | All | 507/1014 | 124 | 112 | 211 | 44 | 16 |
|  |  | CV | 338/676 | 80 | 70 | 147 | 30 | 11 |
|  |  | Holdout | 169/338 | 44 | 42 | 64 | 14 | 5 |
| E4-6 | Gait | All | 317/634 | 82 | 84 | 117 | 28 | 6 |
|  |  | CV | 212/424 | 54 | 53 | 82 | 19 | 4 |
|  |  | Holdout | 105/210 | 28 | 31 | 35 | 9 | 2 |
|  | Balance | All | 320/640 | 76 | 89 | 116 | 32 | 7 |
|  |  | CV | 214/428 | 48 | 53 | 87 | 22 | 4 |
|  |  | Holdout | 106/212 | 28 | 36 | 29 | 10 | 3 |
|  | Voice | All | 446/892 | 112 | 103 | 190 | 34 | 7 |
|  |  | CV | 298/596 | 75 | 71 | 125 | 21 | 6 |
|  |  | Holdout | 148/296 | 37 | 32 | 65 | 13 | 1 |
|  | Tapping | All | 507/1014 | 124 | 112 | 211 | 44 | 16 |
|  |  | CV | 338/676 | 80 | 70 | 147 | 30 | 11 |
|  |  | Holdout | 169/338 | 44 | 42 | 64 | 14 | 5 |

### G. SELECTION OF THE BEST CLASSIFIER DURING THE MAIN EXPERIMENT (NO COMORBIDITIES; MATCHED)

Table 9 shows the classification performance for the four classifiers under consideration for each task.

**Table 9.**
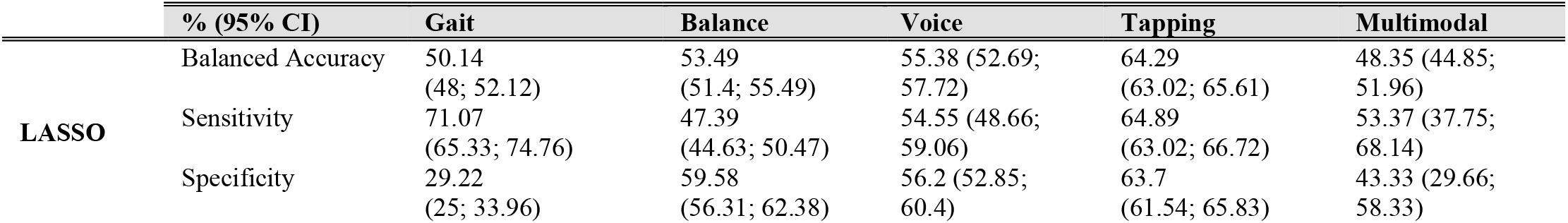
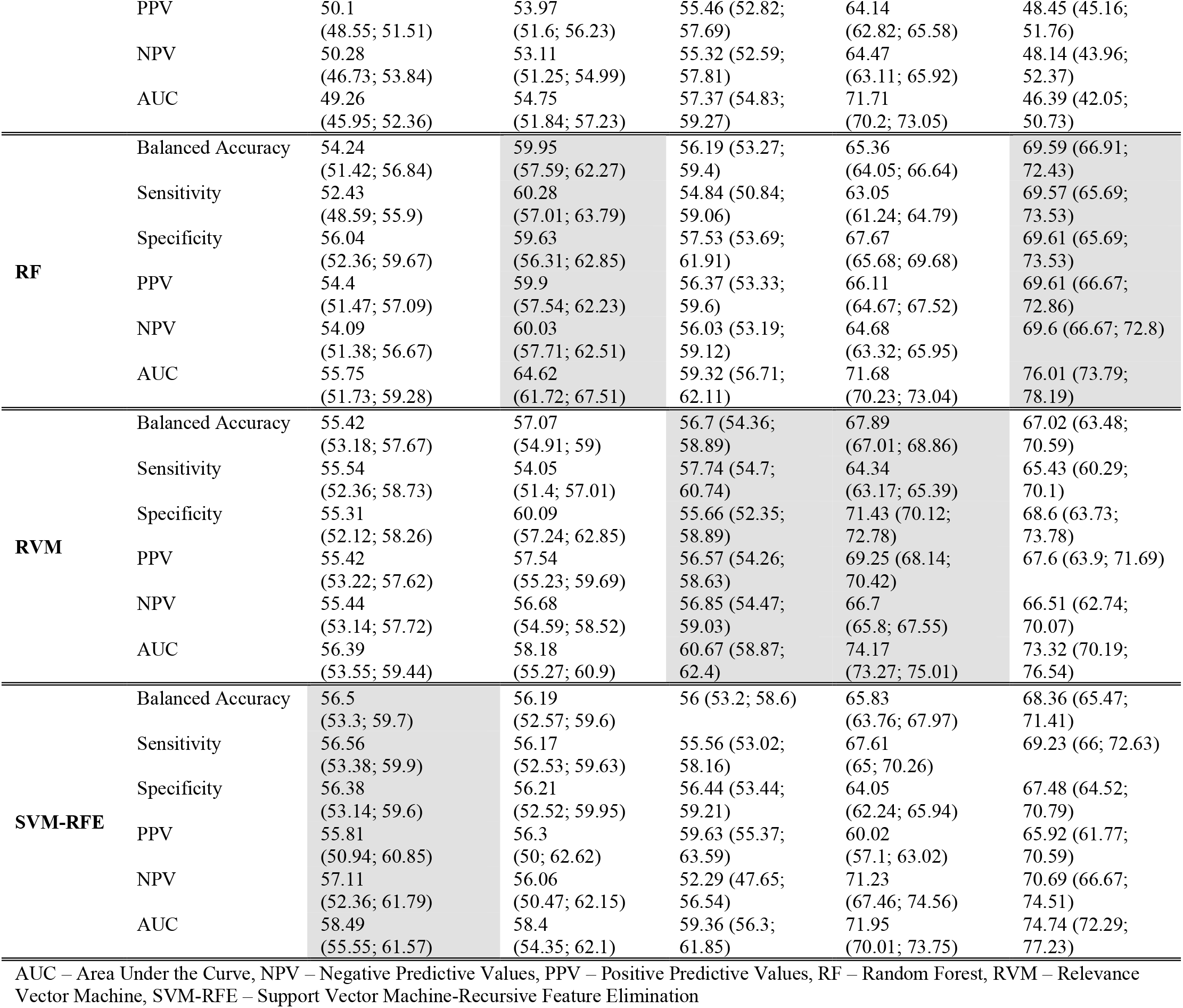
Cross-validation classification performances for each of the tasks (gait, balance, voice, tapping and multimodal features) for four different classifiers.

## APPENDIX II

### SUPPLEMENTARY RESULTS

Table 10-Table 14 summarize the results for each task (gait, balance, voice, tapping) and the combination of all the tasks, for the experiment 1 (all data), experiment 2 (matched data), experiment 3 (no comorbidities and matched data), experiment 4 (no comorbidities, matched, controlled for age), experiment 5 (no comorbidities, matched, controlled for sex) and experiment 6 (no comorbidities, matched, controlled for age and sex).

**Table 10.** Classification performance for the gait task.

|  | Experiment 1 |  |  | Experiment 2 |  |  | Experiment 3 |  |  |
| --- | --- | --- | --- | --- | --- | --- | --- | --- | --- |
|  | CV<br>(95% CI) | Holdout | Chance<br>(95% CI) | CV<br>(95% CI) | Holdout | Chance<br>(95% CI) | CV<br>(95% CI) | Holdout | Chance<br>(95% CI) |
| Balanced accuracy (%) | 56.56<br>(54.34;<br>58.85) | 57.11 | 52.63<br>(51.12;<br>54.21) | 50.33<br>(47.19;<br>53.62) | 54.84 | 50.03<br>(47.13;<br>52.58) | 56.5<br>(53.3;<br>59.7) | 55.71 | 49.92<br>(46.63;<br>53.36) |
| Sensitivity (%) | 52.63<br>(49.52;<br>55.89) | 39.41 | 31.41<br>(26.11;<br>36.45) | 50.34<br>(47.15;<br>53.75) | 50 | 45.21<br>(38.71;<br>50.81) | 56.56<br>(53.38;<br>59.9) | 56.19 | 50.33<br>(43.81;<br>57.14) |
| Specificity (%) | 60.49<br>(59.12;<br>62.02) | 74.81 | 73.86<br>(71.97;<br>76.13) | 50.33<br>(47.15;<br>53.54) | 59.68 | 54.85<br>(54.35;<br>55.56) | 56.38<br>(53.14;<br>59.6) | 55.24 | 49.52<br>(49.45;<br>49.58) |
| PPV (%) | 38.22<br>(34.64;<br>42.02) | 54.79 | 50<br>(50; 50) | 49.68<br>(45.38;<br>54.22) | 55.36 | 50<br>(50; 50) | 55.81<br>(50.94;<br>60.85) | 55.66 | 50<br>(50; 50) |
| NPV (%) | 73.32<br>(70.1;<br>76.19) | 61.44 | 56.35<br>(52.98;<br>59.56) | 50.98<br>(46.19;<br>55.82) | 54.41 | 50.05<br>(44.12;<br>55.15) | 57.11<br>(52.36;<br>61.79) | 55.77 | 49.85<br>(43.27;<br>56.73) |
| AUC (%) | 59.45<br>(57.41;<br>61.38) | 59.88 | 50.03<br>(44.42;<br>55.17) | 50.98<br>(47.84;<br>54.13) | 56.5 | 50.07<br>(42.9; 56.99) | 58.49<br>(55.55;<br>61.57) | 55.85 | 49.95<br>(41.48;<br>57.74) |

|  | Experiment 4 |  |  | Experiment 5 |  |  | Experiment 6 |  |  |
| --- | --- | --- | --- | --- | --- | --- | --- | --- | --- |
|  | CV<br>(95% CI) | Holdout | Chance<br>(95% CI) | CV<br>(95% CI) | Holdout | Chance<br>(95% CI) | CV<br>(95% CI) | Holdout | Chance<br>(95% CI) |
| Balanced accuracy (%) | 56.48<br>(53.3;<br>59.46) | 54.76 | 49.97<br>(46.7; 53.31) | 56.4<br>(53.07;<br>59.68) | 56.19 | 50<br>(46.43;<br>53.57) | 56.65<br>(53.3;<br>59.91) | 53.81 | 50.05<br>(46.85;<br>53.19) |
| Sensitivity (%) | 56.56<br>(53.24;<br>59.66) | 54.29 | 49.46<br>(42.86;<br>56.19) | 56.46<br>(53.08;<br>59.95) | 56.19 | 49.99<br>(42.86;<br>57.14) | 56.71<br>(53.32;<br>60.05) | 51.43 | 47.71<br>(40.95;<br>54.29) |
| Specificity (%) | 56.39<br>(53.16;<br>59.43) | 55.24 | 50.48<br>(50.42;<br>50.55) | 56.35<br>(52.93;<br>59.62) | 56.19 | 50<br>(50; 50) | 56.58<br>(53.24;<br>59.91) | 56.19 | 52.39<br>(52.1; 52.75) |
| PPV (%) | 55.82<br>(51.42;<br>60.38) | 54.81 | 50<br>(50; 50) | 56<br>(50.94;<br>60.38) | 56.19 | 50<br>(50; 50) | 56.12<br>(51.42;<br>60.38) | 54 | 50<br>(50; 50) |
| NPV (%) | 57.12<br>(52.36;<br>61.79) | 54.72 | 49.94<br>(43.4; 56.6) | 56.82<br>(52.36;<br>61.32) | 56.19 | 49.99<br>(42.86;<br>57.14) | 57.15<br>(52.83;<br>61.79) | 53.64 | 50.08<br>(43.64;<br>56.36) |
| AUC (%) | 58.51<br>(55.51;<br>61.3) | 56.25 | 49.99<br>(41.91;<br>58.25) | 58.2<br>(54.83;<br>61.24) | 56.1 | 50.02<br>(42.65;<br>58.05) | 58.33<br>(55.25;<br>61.52) | 55.99 | 50.07 (42.16;<br>57.99) |

AUC – Area Under the Curve, CV – Cross-Validation, NPV – Negative Predictive Values, PPV – Positive Predictive Values

**Table 11.**
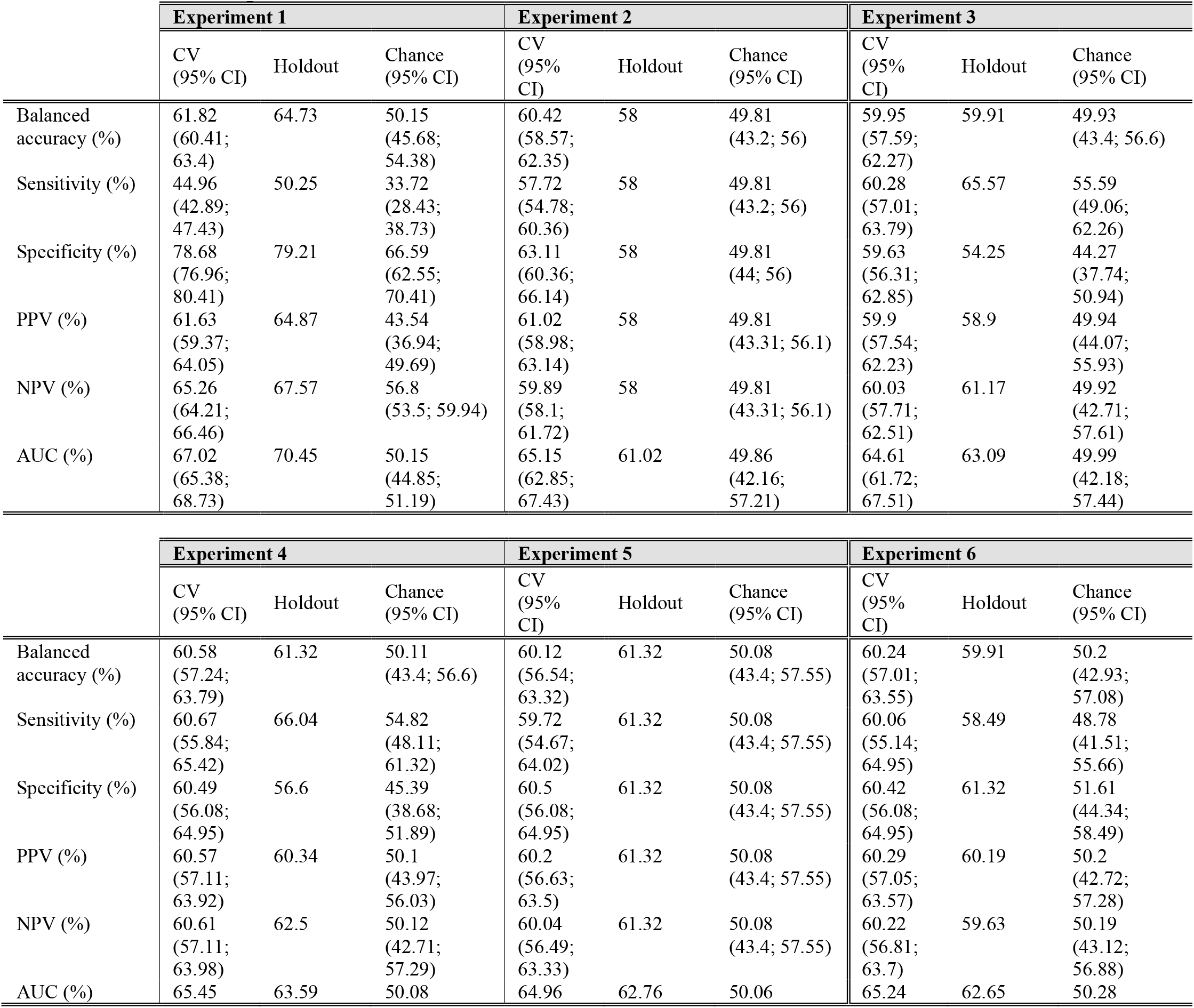
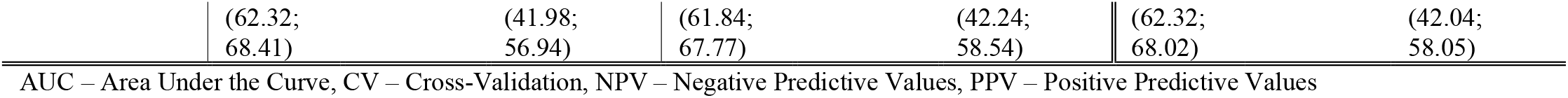
Classification performance for the balance task.

**Table 12.** Classification performance for the voice task.

|  | Experiment 1 |  |  | Experiment 2 |  |  | Experiment 3 |  |  |
| --- | --- | --- | --- | --- | --- | --- | --- | --- | --- |
|  | CV<br>(95% CI) | Holdout | Chance<br>(95% CI) | CV<br>(95% CI) | Holdout | Chance<br>(95% CI) | CV<br>(95% CI) | Holdout | Chance<br>(95% CI) |
| Balanced accuracy (%) | 62.49<br>(61.33;<br>63.62) | 60.41 | 49.99 (46.46;<br>53.5) | 53.94<br>(51.48;<br>56.18) | 59.83 | 50.05 (44.66;<br>55.34) | 56.7<br>(54.36;<br>58.89) | 53.04 | 49.98 (43.58;<br>55.74) |
| Sensitivity (%) | 46.54<br>(44.63;<br>48.49) | 44.44 | 32.25 (28.11;<br>36.36) | 50.43<br>(47.05;<br>53.65) | 56.74 | 46.96 (41.57;<br>52.25) | 57.74<br>(54.7;<br>60.74) | 52.03 | 48.97 (42.57;<br>54.73) |
| Specificity (%) | 78.43<br>(77.15;<br>79.71) | 76.37 | 67.73 (64.8;<br>70.64) | 57.45<br>(53.93;<br>60.67) | 62.92 | 53.14 (47.75;<br>58.43) | 55.66<br>(52.35;<br>58.89) | 54.05 | 50.99 (44.6;<br>56.76) |
| PPV (%) | 60.55<br>(58.79;<br>62.19) | 57.14 | 41.47 (36.15;<br>46.75) | 54.24<br>(51.62;<br>56.67) | 60.48 | 50.05 (44.31;<br>55.69) | 56.57<br>(54.26;<br>58.63) | 53.1 | 49.98 (43.45;<br>55.86) |
| NPV (%) | 67.35<br>(66.5;<br>68.19) | 65.98 | 58.51 (55.98;<br>61.03) | 53.68<br>(51.36;<br>55.76) | 59.26 | 50.04 (44.97;<br>55.03) | 56.85<br>(54.47;<br>50.03) | 52.98 | 49.98 (43.71;<br>55.63) |
| AUC (%) | 68.99<br>(68.14;<br>69.79) | 66.95 | 49.93 (45.49;<br>54.03) | 55.5<br>(53.36;<br>57.6) | 62.48 | 50.14 (44.18;<br>56.09) | 60.67<br>(58.87;<br>62.4) | 54.99 | 50.01 (43.02;<br>56.65) |

|  | Experiment 4 |  |  | Experiment 5 |  |  | Experiment 6 |  |  |
| --- | --- | --- | --- | --- | --- | --- | --- | --- | --- |
|  | CV<br>(95% CI) | Holdout | Chance<br>(95% CI) | CV<br>(95% CI) | Holdout | Chance<br>(95% CI) | CV<br>(95% CI) | Holdout | Chance<br>(95% CI) |
| Balanced accuracy (%) | 56.85<br>54.7;<br>59.06) | 58.11 | 49.98 (44.6;<br>56.08) | 59.99<br>(57.72;<br>62.08) | 60.14 | 49.82 (43.92;<br>55.41) | 59.15<br>(57.05;<br>61.24) | 59.12 | 50.15 (43.58;<br>55.74) |
| Sensitivity (%) | 56.64<br>(53.69;<br>59.56) | 53.38 | 45.25 (39.87;<br>51.35) | 59.55<br>(56.38;<br>62.42) | 53.38 | 43.06 (37.16;<br>48.65) | 58.83<br>(56.04;<br>61.41) | 58.78 | 49.82 (43.24;<br>55.41) |
| Specificity (%) | 57.06<br>(54.03;<br>60.07) | 62.84 | 54.71 (49.32;<br>60.81) | 60.43<br>(57.38;<br>63.42) | 66.89 | 56.58 (50.68;<br>62.16) | 59.46<br>(56.71;<br>62.42) | 59.46 | 50.49 (43.92;<br>56.08) |
| PPV (%) | 56.88<br>(54.59;<br>59.25) | 58.96 | 49.98 (44.03;<br>56.72) | 60.08<br>(57.76;<br>62.2) | 61.72 | 49.79 (42.97;<br>56.25) | 59.21<br>(57;<br>61.36) | 59.18 | 50.15 (43.54;<br>55.78) |
| NPV (%) | 56.82<br>(54.64;<br>59.06) | 57.41 | 49.98 (45.06;<br>55.56) | 59.91<br>(57.69;<br>62) | 58.93 | 49.84 (44.64;<br>54.76) | 59.09<br>(57.02;<br>61.23) | 59.06 | 50.15 (43.62;<br>55.71) |
| AUC (%) | 58.44<br>(56.37;<br>60.38) | 59.13 | 50.03 (43.7;<br>56.47) | 63.3<br>(61.25;<br>65.05) | 61.64 | 49.85 (43.28;<br>56.58) | 61.72<br>(59.65;<br>63.53) | 63.26 | 50.25 (43.32;<br>57.15) |

AUC – Area Under the Curve, CV – Cross-Validation, NPV – Negative Predictive Values, PPV – Positive Predictive Values

**Table 13.**
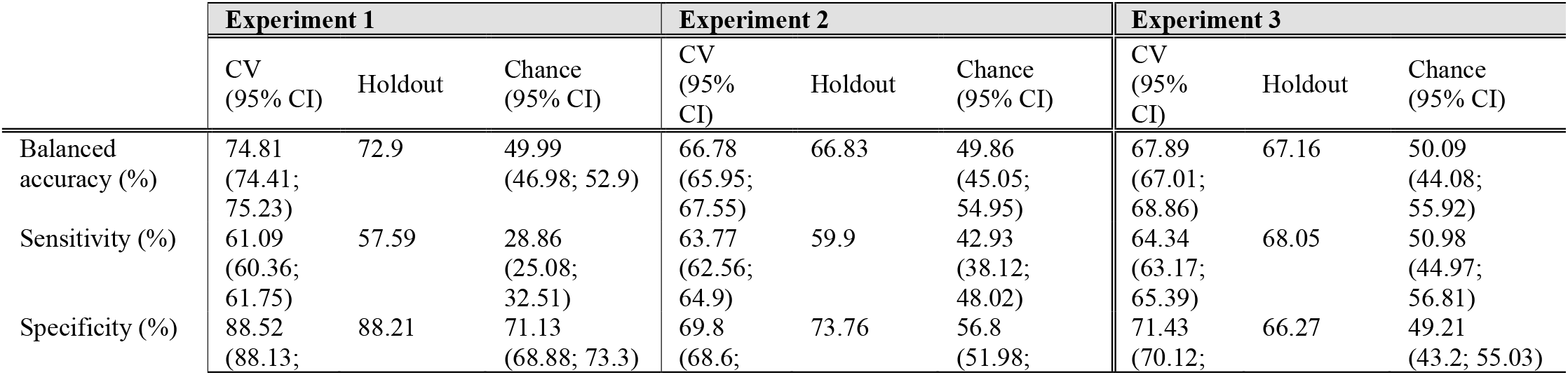
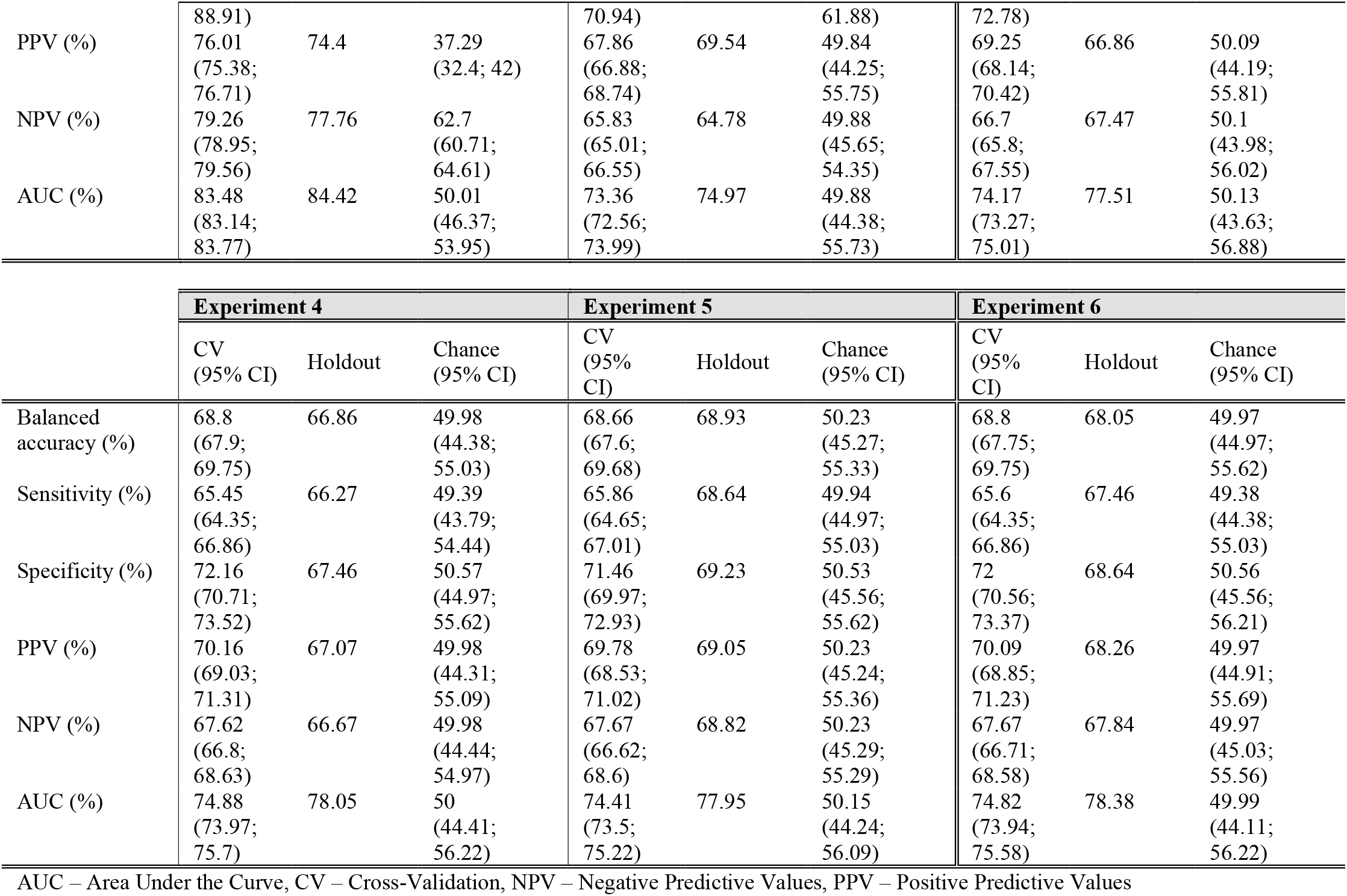
Classification performance for the tapping task.

**Table 14.**
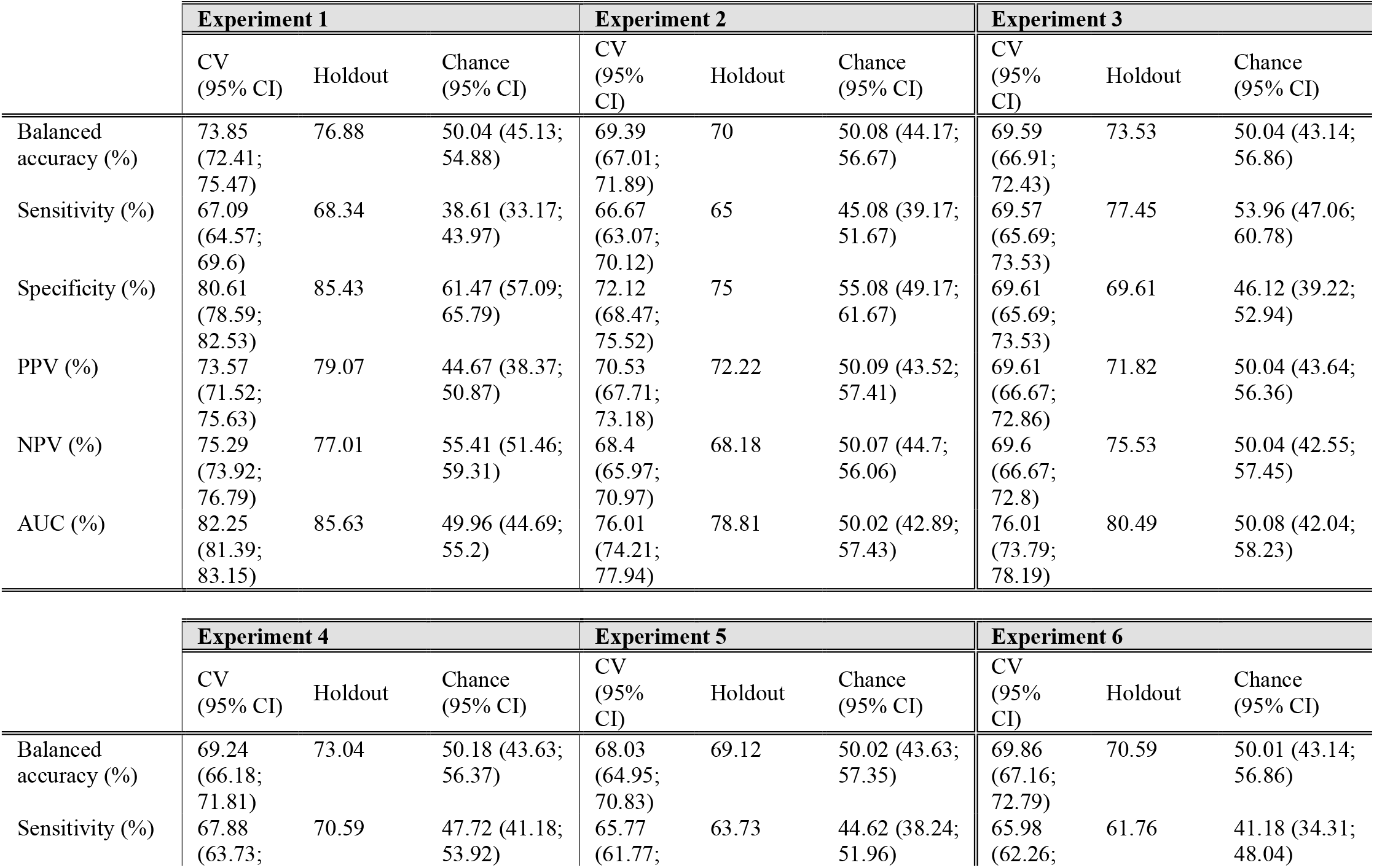
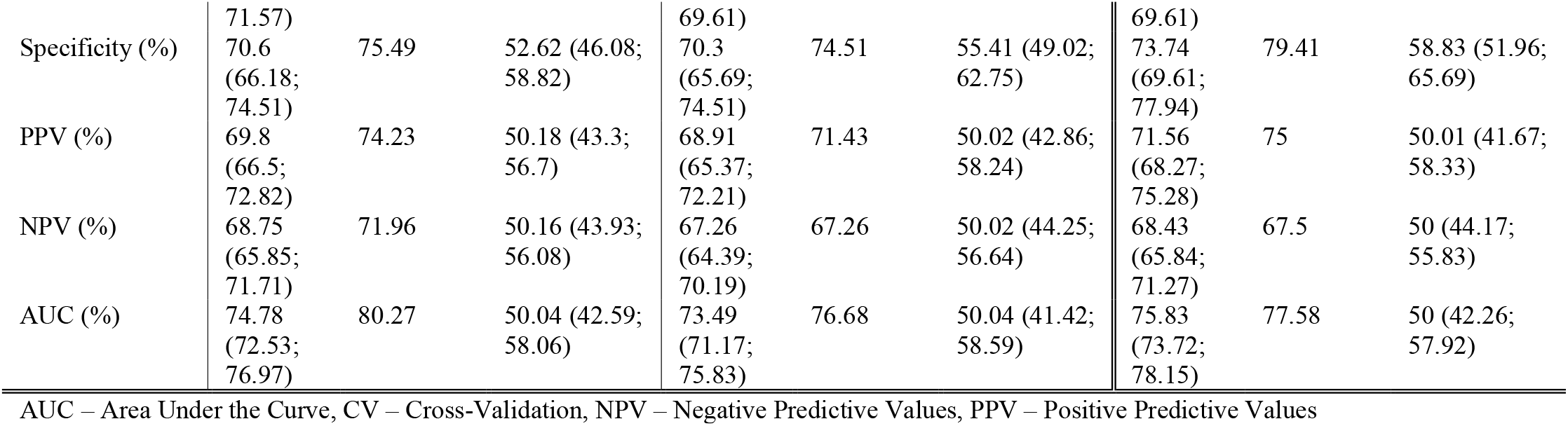
Classification performance for the multimodal features.

#### A. ADDITIONAL EXPERIMENTS

Our results may differ to those in the current literature using the mPower dataset since we follow different approaches. To explain these discrepancies and compare with the literature, we included two additional experiments including all data without trimming for age range and all data including both age and sex as features in the analyses (Table 15). Classification performances for both additional experiments for each tasks are summarized in Table 16-Table 20.

**Table 15.**
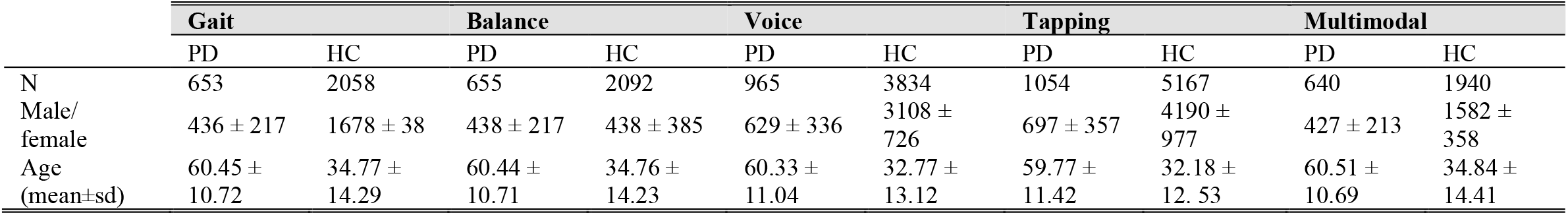
Demographics for PD and HC subjects including all data.

**Table 16.** Classification performance for the gait task.

|  | Additional experiment:<br>All data |  |  | Additional experiment:<br>All data + age + gender |  |  |
| --- | --- | --- | --- | --- | --- | --- |
|  | CV<br>(95% CI) | Holdout | Chance<br>(95% CI) | CV<br>(95% CI) | Holdout | Chance<br>(95% CI) |
| Balanced<br>accuracy (%) | 50.95<br>(50.29;<br>51.63) | 51.55 | 50.01 (49.13;<br>50.95) | 57.2<br>(55.81;<br>58.68) | 58.25 | 50.01 (48.54;<br>51.57) |
| Sensitivity (%) | 2.74<br>(1.61;<br>4.13) | 3.69 | 1.35 (0;<br>2.77) | 16.19<br>(13.3;<br>19.27) | 17.51 | 5 (2.77;<br>7.37) |
| Specificity (%) | 99.16<br>(98.8;<br>99.6) | 99.42 | 98.68 (98.25;<br>99.13) | 98.21<br>(97.63;<br>98.76) | 98.98 | 95.02 (94.32;<br>95.77) |
| PPV (%) | 50.8<br>(33.33;<br>66.67) | 66.67 | 24.34 (0; 50) | 74.26<br>(68.28;<br>80.66) | 84.44 | 24.12 (13.33;<br>35.56) |
| NPV (%) | 76.24<br>(75.99;<br>76.49) | 76.54 | 75.97 (75.65;<br>76.32) | 78.67<br>(78.11;<br>79.26) | 79.14 | 75.97 (75.41;<br>76.57) |
| AUC (%) | 66.08<br>(64.67;<br>67.52) | 67.81 | 49.93 (45.49;<br>54.46) | 86.49<br>(85.66;<br>87.32) | 88.43 | 50.01 (45.45;<br>54.51) |

**Table 17.**
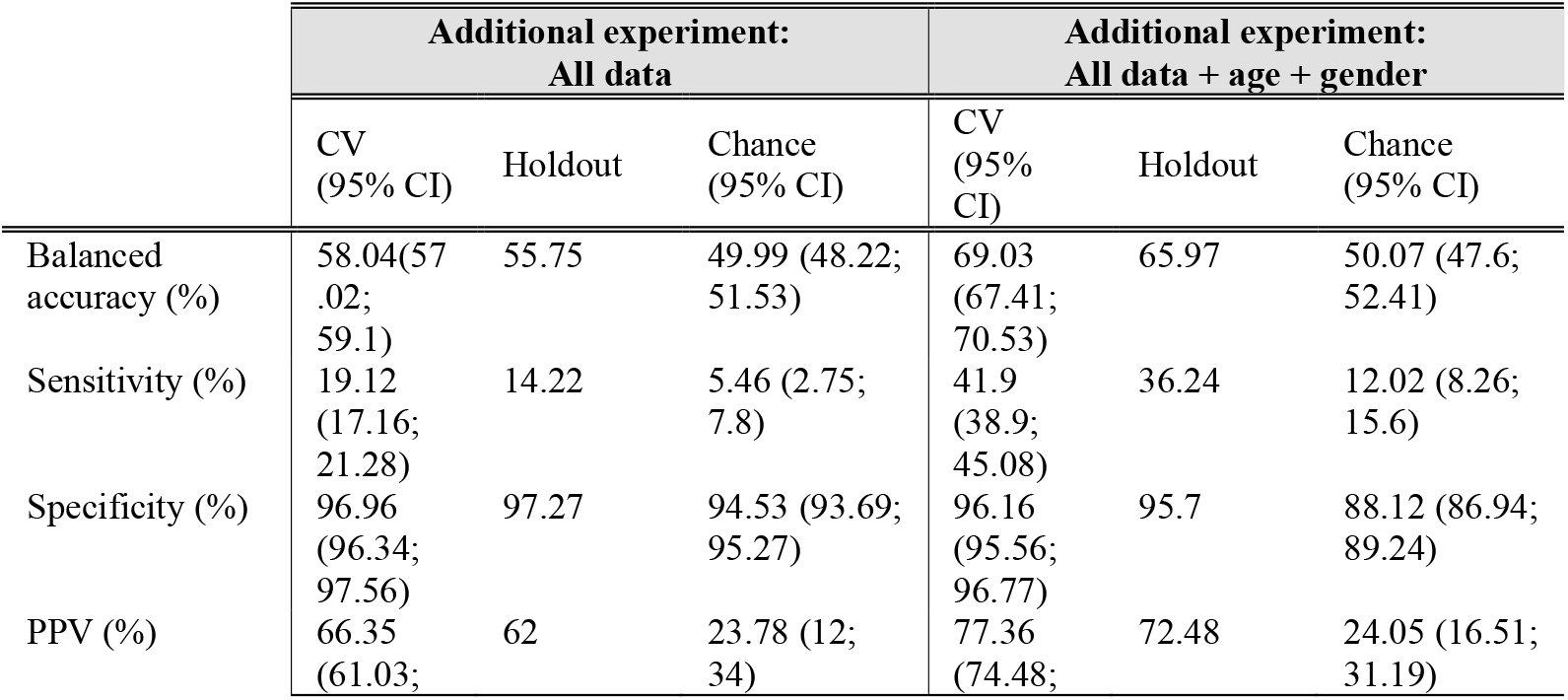
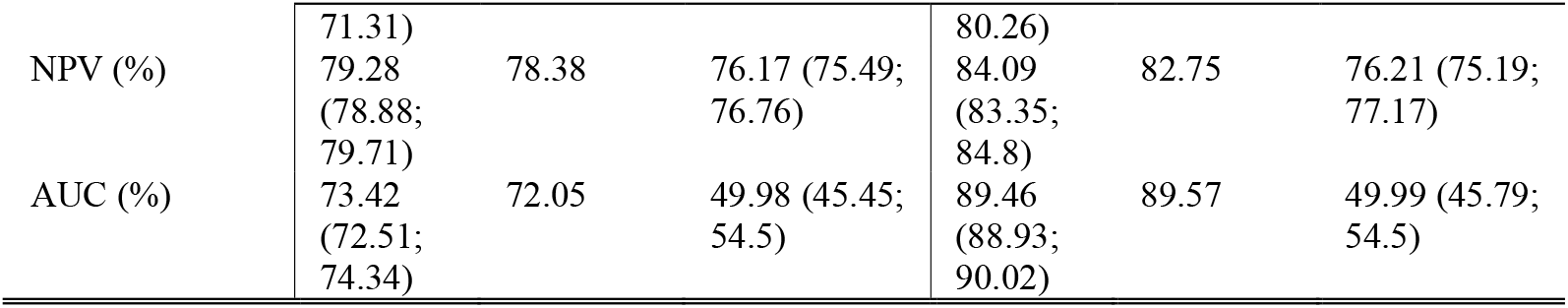
Classification performance for the balance task.

**Table 18.** Classification performance for the voice task.

|  | Additional experiment:<br>All data |  |  | Additional experiment:<br>All data + age + gender |  |  |
| --- | --- | --- | --- | --- | --- | --- |
|  | CV<br>(95% CI) | Holdout | Chance<br>(95% CI) | CV<br>(95% CI) | Holdout | Chance<br>(95% CI) |
| Balanced accuracy (%) | 52.91<br>(52.13;<br>53.68) | 52.57 | 50.03 (49.26;<br>51.01) | 73.12<br>(71.9;<br>74.35) | 74.6 | 50 (48.09;<br>52.19) |
| Sensitivity (%) | 7.59<br>(6.06;<br>9.16) | 6.23 | 2.17 (0.94;<br>3.74) | 50.68<br>(48.29;<br>53.11) | 53.89 | 14.58 (11.53;<br>18.07) |
| Specificity (%) | 98.24<br>(97.93;<br>98.55) | 98.9 | 97.89 (97.57;<br>98.28) | 95.55<br>(95.11;<br>96.01) | 95.31 | 85.43 (84.66;<br>86.31) |
| PPV (%) | 52.05<br>(45.24;<br>58.7) | 58.82 | 20.49 (8.82;<br>35.29) | 74.18<br>(72.28;<br>76.2) | 74.25 | 20.08 (15.88;<br>24.89) |
| NPV (%) | 80.84<br>(80.58;<br>81.1) | 80.77 | 79.93 (79.68;<br>80.26) | 88.49<br>(88; 89) | 89.17 | 79.93 (79.21;<br>80.75) |
| AUC (%) | 73.23<br>(72.43;<br>74.01) | 74.91 | 50.09 (46.63;<br>53.52) | 91.39<br>(91.05;<br>91.72) | 91.16 | 49.99 (46.49;<br>53.58) |

**Table 19.**
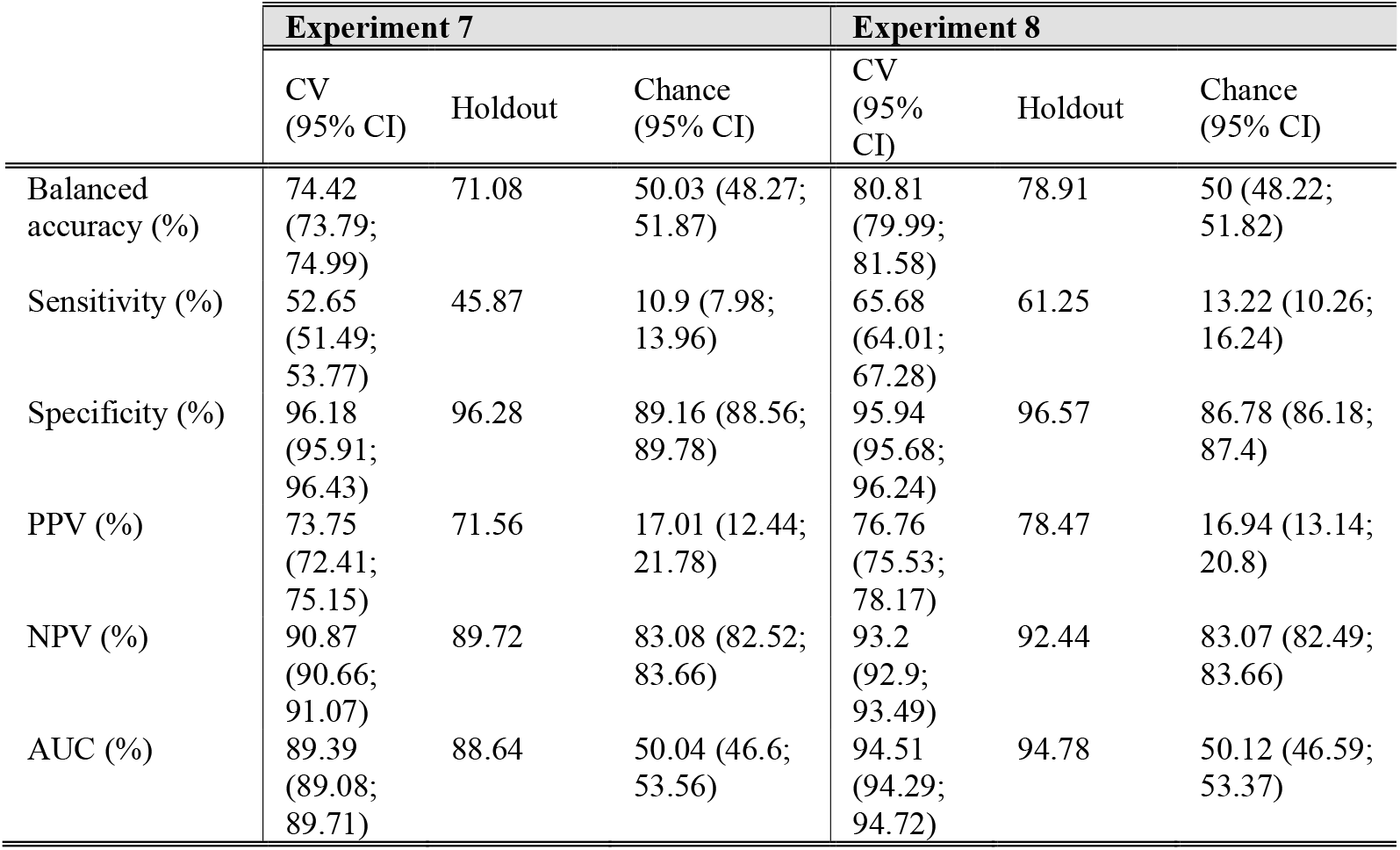
Classification performance for the tapping task.

**Table 20.**
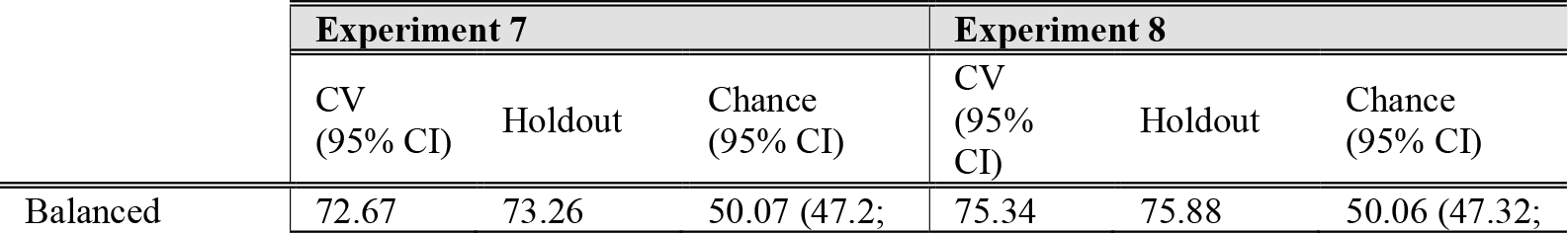
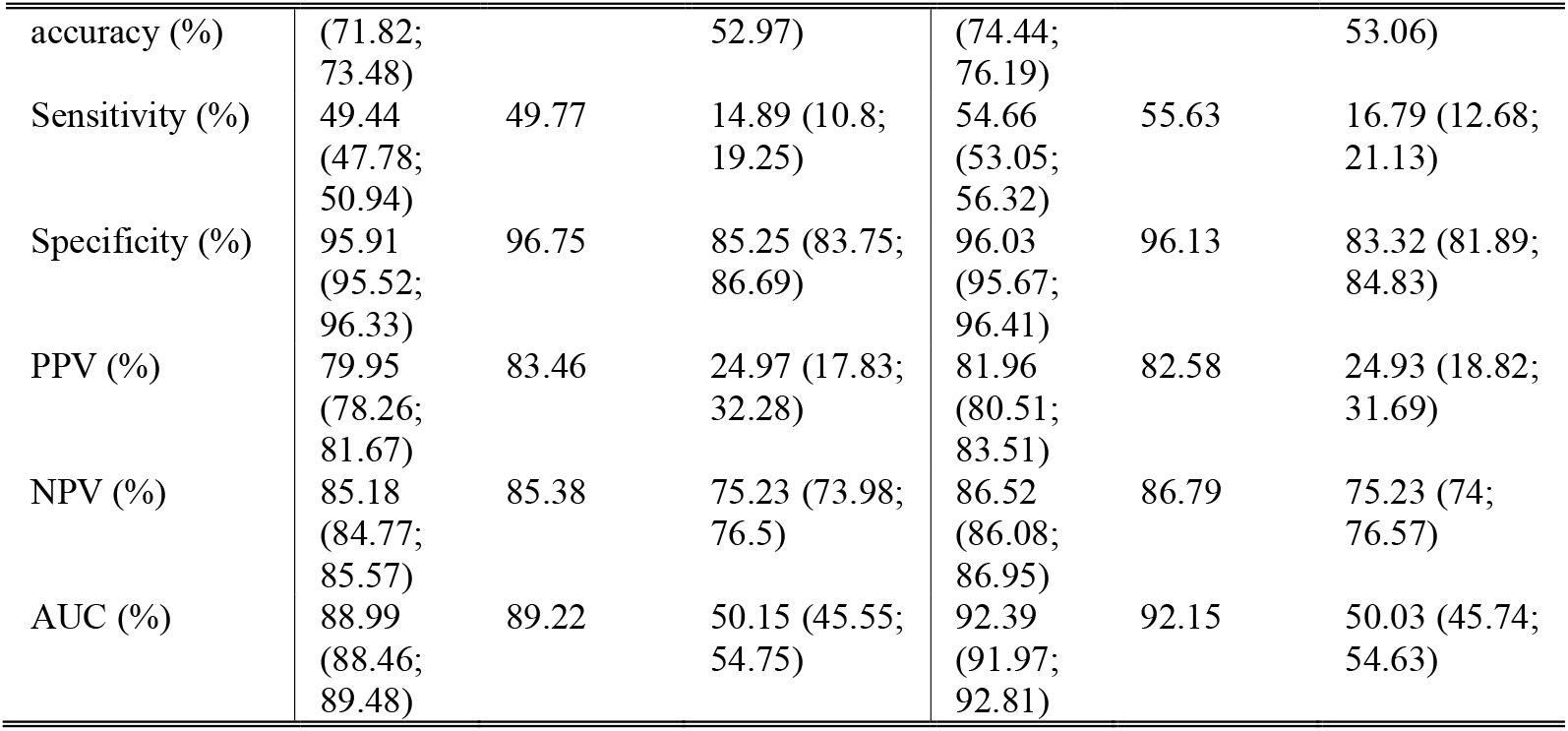
Classification performance for the multimodal features.

## ACKNOWLEDGMENT

Data used in this study were contributed by users of the Parkinson m-Power mobile application as part of the m-Power study developed by Sage Bionetworks and described in Synapse [doi:10.7303/syn4993293].

## AUTHOR CONTRIBUTION

MG performed the overall analyses and wrote manuscript. JD designed the overall study and contributed to writing the manuscript. KRP, SBE and MSF provided input on the analyses. All authors contributed to interpretation of results, reviewed and commented on the manuscript.

## AUTHOR DECLARATION

The access to the Mpower data was granted after registration in the Synapse system, signing an oath, submitting an Intended Data Use Statement and accepting data-specific Conditions. All appropriate institutional forms have been archived.

